# A high-resolution genomic and phenotypic analysis of resistance evolution of an *Escherichia coli* strain from a critical care patient treated with piperacillin/tazobactam

**DOI:** 10.1101/2024.01.15.24301199

**Authors:** Alice J. Fraser, Robert Ball, Daire Cantillon, Laura E. Brettell, Fabrice E. Graf, John T. Munnoch, Paul A. Hoskisson, Joseph M. Lewis, Jon J. van Aartsen, Christopher M. Parry, Eva Heinz, Thomas Edwards

**Author notes:** Contributed equally (Eva Heinz, Thomas Edwards).

## Abstract

Resistance to the β-lactam/β-lactamase inhibitor (BL/BLI) combination antibiotic piperacillin/tazobactam (TZP) predominantly occurs via β-lactamase enzymes also leading to resistance to third-generation cephalosporins (3GCs). However, if β-lactamases inactive against 3GCs and inhibited by tazobactam are expressed at high levels leading to enzyme hyperproduction, the surplus enzyme escapes inhibition by tazobactam and inactivates the antibiotic piperacillin. Understanding this mechanism is clinically relevant as enzyme hyperproduction can emerge upon antibiotic administration, resulting in treatment failure despite initial resistance profiles supporting TZP use.

We report the identification of an *Escherichia coli* isolate that developed resistance to TZP during patient treatment. Our whole genome sequencing (WGS) analyses show that TZP resistance evolved via IS*26*-mediated duplication of a *bla*TEM-1 containing gene cassette on a plasmid, resulting in hyperproduction of TEM-1 β-lactamase. We demonstrate that ten copies of *bla*TEM-1 induce resistance greater than 32-times the MIC and exposure to TZP further increases amplification of *bla*TEM-1. Furthermore, in the absence of TZP, gene copy number of *IS26* and *bla*TEM-1 remains stable over five days, despite a 48,205 bp genome size increase compared to the pre-amplification isolate. We additionally detect phenotypic changes that might indicate host adaptation potentially linked to the additional genes in the amplified cassette.

Our analysis advances the understanding of infections caused by isolates evolving β-lactamase hyperproduction, which represent a complex problem in both detection and treatment. As 40% of antibiotics active against WHO priority pathogens in the pre-clinical pipeline are BL/BLI combinations further investigations are of urgent concern.

Importance

We investigated an *Escherichia coli* strain obtained from the bloodstream of a hospitalised patient, that evolved resistance against the antimicrobials initially used as empirical treatment. Comparing the whole-genome sequences of the susceptible isolate with the evolved, resistant isolate showed duplications of the only encoded β-lactamase gene, *bla*TEM-1, resulting in increased enzyme production and resistance to TZP, a commonly prescribed BL/BLI combination antimicrobial. Despite the additional energy needed for increased enzyme production and retaining the additional copies of duplicated genes, we did not find growth differences under standard laboratory conditions and when using a high-throughput metabolic screen. We did however identify phenotypic changes that indicate host adaptation and mirror phenotypic changes observed in other species of opportunistic bacterial pathogens. In summary our findings highlight that BL/BLI combinations can lead to rapid within-patient evolution of antimicrobial resistance, which is of high relevance when considering the implementation of newly developed drugs, many of which belong to the BL/BLI class.

## Introduction

The spectrum of clinical disease caused by *Escherichia coli* infections ranges from uncomplicated localised infections, such as those affecting the lower urinary tract, to disseminated and serious infections, including bacteraemia. In the case of infections which result in patient hospitalisation, antimicrobials belonging to the β-lactam class are often used as an empirical treatment, primarily due to their broad spectrum of activity and safe use (1). Cephalosporins, especially third-generation cephalosporins (3GCS), such as cefotaxime, ceftazidime and ceftriaxone are β-lactams suitable for treating suspected sepsis, pneumonia and meningitis (2). In cases where the patient is critically ill or a resistance to 3GCs is likely, carbapenems such as ertapenem, imipenem or meropenem may be used. These antimicrobials are generally considered antimicrobials of “last resort” beyond which treatments options are limited (3). Pathogenic *E. coli* strains are now often resistant to early generations of β-lactam drugs, primarily due to the production of β-lactamase enzymes, acquired via mobile genetic elements. Extended-spectrum β-lactamases (ESBLs), which have increased hydrolytic capability and can break down 3GCs, are increasingly prevalent in *E. coli* globally and lead to treatment failures of common first- and second-line agents. Collectively these put our ability to provide effective treatments at risk and in 2019 over 250,000 deaths worldwide were attributed to antimicrobial resistant *E. coli* infections (4).

To address the increase in infections caused by drug-resistant, β-lactamase-producing *E. coli* strains, the combination of a β-lactamase inhibitor with a β-lactam antibiotic is a common strategy used to restore clinical efficacy. The hydrolytic activity of the β-lactamase is countered by the binding of the inhibitor to the enzyme, thus leaving the antibiotic free to exert its bactericidal activity and render isolates *de facto* susceptible.

Piperacillin/tazobactam (TZP) is a clinically important β-lactam/β-lactamase-inhibitor (BL/BLI) combination therapy, indicated for the first-line empirical treatment of serious infections including those escalating to bacteraemia and caused by *E. coli* (5). Tazobactam, a “suicide inhibitor”, irreversibly binds to the β-lactamase present, rendering it inactive and thus restoring the activity of piperacillin, a penicillin β-lactam class antibiotic. Tazobactam has *in vitro* activity against Ambler class A and Ambler class C β-lactamases, which includes AmpC, TEM, SHV and CTX-M enzymes, is well tolerated and, combined with piperacillin, has broad-spectrum activity against both Gram-negative and Gram-positive pathogens (6).

The introduction of TZP as a treatment option however has rapidly been followed by observations of resistant strains. Mechanisms which reduce susceptibility to TZP may be independent of β-lactamase production, such as increased drug efflux due to efflux pump overexpression (7); decreased influx, due to porin downregulation (8); and mutations in penicillin binding proteins (PBPs), leading to reduced binding of the penicillin (9).

Alternatively, production of multiple β-lactamases (10), which can enable the bacteria to overcome the inhibitory effects of tazobactam via saturation of the inhibitor; or production of carbapenemases (11), to which tazobactam lacks activity, may permit β-lactamase hydrolysis of the penicillin.

In some strains selective resistance to TZP occurs, whilst susceptibility to 3GCs is retained. This can be induced by hyperproduction of enzymes which would otherwise be blocked by tazobactam, but the increased production means excess enzymes can thus escape the inhibitor and function as normal. The other cause of this phenotype can be mutations in enzymes which would otherwise not induce TZP resistance. In *E. coli* different resistance mechanisms have evolved which induce hyperproduction of TEM and SHV enzymes. These include mutations in promoter regions (12), gene duplication events (13–15), and increase in plasmid copy number (16). Alternatively, the acquisition of CTX-M-15 enzyme variants exhibiting the S133G mutation can induce selective resistance to TZP (17). The TZP-resistant/3GC-susceptible (TZP-R/3GC-S) phenotype is not restricted to particular *E. coli* clones and has been reported in a wide range of sequence types (18). Selecting an appropriate treatment for these strains can be challenging. Whilst treatment with a 3GC could be effective and would reduce exposure to antimicrobials of last-resort; TZP treatment failure can also be indicative of ESBL or high-level AmpC production (19) . Carbapenem use may therefore be considered by some clinicians as a safer option to ensure successful treatment is not further delayed. (20).

Here we report the characterisation of a clinical *E. coli* strain isolated from blood culture, which evolved resistance during treatment of a patient with TZP, resulting in the emergence of the previously described TZP-R/3GC-S phenotype. Using whole genome sequencing and subsequent bioinformatic analyses, combined with *in vitro* laboratory investigations, we describe the cause of the resistance phenotype; an extensive gene amplification of *bla*TEM-1 resulting in β-lactamase hyperproduction. We also highlight that whilst successful treatment included carbapenems, the use of 3GC may have also been suitable and thus prevented the use of an antimicrobial of last resort.

## Methods

### Ethics

Both *E. coli* isolates used in this study were collected from the Royal Liverpool University Hospital (RLUH; Liverpool, UK), as part of routine clinical diagnostic procedures. Informed consent was obtained from the patient to publish case details and carry out experimental work, and the study was approved by Liverpool School of Tropical Medicine Research Ethics Committee (Study number 22-074). Isolates were retrieved from the hospital biobank, under a Material Transfer Agreement between Liverpool Clinical Laboratories (LCL) and LSTM.

### Bacteria isolates, storage and media

Clinical *E. coli* isolates were grown from blood cultures performed at LCL on the day of admission and day five post-admission. Isolates were stored in glycerol broth in Microbank tubes (Pro-lab Diagnostics, U.K.) at -80 °C and resurrected from glycerol stocks on Luria-Bertani (LB) agar (Sigma, U.K.) at 37 °C for 18 hours. Single colonies were then picked and used for subsequent experiments.

Control strains of *Bacteroides thetaiotaomicron* (VPI-5482) and *Pseudomonas aeruginosa* (NCTC 13437) were used as positive controls in our anaerobic growth assay and for the assay assessing biofilm formation, respectively.

Both were resurrected from glycerol stocks and grown on brain heart infusion (BHI) agar (Sigma) supplemented with 5 mg/L of hemin (Sigma) and LB agar, respectively.

### Antimicrobial susceptibility testing

Initial antimicrobial susceptibility testing (AST) at LCL was performed using the disk diffusion method according to EUCAST guidelines. Susceptibility to 3GCs was inferred from cefpodoxime. Upon receipt, we verified the TZP phenotypes where the susceptible isolate was susceptible to TZP (MIC 1-2/4 μg/ml) and the resistant isolate was resistant (MIC 256-512/4 μg/ml) to TZP via broth microdilution, according the guidelines as set out by EUCAST(21), which states the clinical breakpoint as 8/4 μg/ml (22). Piperacillin and tazobactam (both Sigma, UK) were solubilised in DMSO (Sigma) to a stock concentration of 10 mg/ml and then sterilised through a 0.22 μm polyethersulfone filter unit (Milipore, USA). Triplicate liquid cultures were grown from the selection of three distinct colonies, and each liquid culture was then tested in triplicate.

### Extraction of DNA for whole genome sequencing and quantitative PCR

Single colonies were transferred to 10 ml of LB broth (Sigma, UK), and grown overnight at 37°C, 200 RPM. Long fragments of genomic DNA, used for Oxford Nanopore Technologies (ONT) sequencing, were extracted using the Masterpure™ Complete DNA and RNA Purification Kit (Lucigen, U.K.), following the manufacturer’s instructions for the purification of DNA from cell samples. Short fragment DNA, used for Illumina sequencing and qPCR, were extracted using the DNeasy™ Blood and Tissue Kit (Qiagen, Germany), following the protocol for Gram-negative Bacteria. The quality and size of the short-fragment DNA used for Illumina sequencing was assessed using the TapeStation (4150) system and the DNA ScreenTape Kit (Agilent, USA). All genomic DNA used for sequencing was quantified using the Qubit Fluorometer with the dsDNA BR Kit (Invitrogen, USA).

### Whole genome sequencing

Short-read sequencing was performed by Microbes NG (Birmingham, UK). Genomic DNA libraries are prepared using the Nextera XT Library Prep Kit (Illumina, USA) following the manufacturer’s protocol with the following modifications: input DNA is increased 2-fold, and PCR elongation time is increased to 45 seconds. DNA quantification and library preparation are carried out on a Hamilton Microlab STAR automated liquid handling system (Hamilton Bonaduz AG, Switzerland). Libraries are sequenced on an lllumina NovaSeq 6000 using a 250 bp paired end protocol.

Long-read sequencing was performed on a MinION MK1B sequencing device (ONT, U.K.). Library preparation was carried out according to the manufacturers protocol, using the ligation sequencing kit (SQK-LSK109) and Native Barcoding Expansion Kits (EXP-NBD104; all ONT). Sequencing was carried out using a FLOW-MIN106 R9.4.1 flow cell (ONT), the libraries were loaded as a pool containing 10 sample libraries in total. Illumina and ONT reads can be found at the Sequence Read Archive bioproject ID: PRJNA1061590.

### Assemblies

The generated fastq files then underwent trimming using Trimmomatic (v0.39) (23), with a sliding window quality cut-off value of Q20 and were quality assessed using FASTQC (v0.11.9) (24).

Basecalling and de-multiplexing of long reads was performed with Guppy (v5.0.7) (25), using the super-accurate model for basecalling. Long read sequences were then filtered using Filtlong (v0.2.1) (26) and adapters trimmed using Porechop (v0.2.4) (27).

Long-read-first hybrid assemblies were produced using Unicycler (v0.5.0) (28), with the Miniasm+Racon pipeline, these were then visualized using Bandage (v0.8.1) (29), before being polished with long reads using Medaka (v1.5.0) (30). Assemblies then underwent polishing with short reads using Polypolish (v0.5.0) (31) and NextPolish (1.4.1) (32). Finally, polished assemblies were quality checked, using the bwa-mem (v0.7.17) (33) and samtools (v1.9) (34) to map reads back to the consensus assembly sequence. These were then visualised using Artemis (v18.2.0) (35) and the Artemis comparison tool(v18.2.0) (36).

### Genome analyses

Polished assemblies were annotated using PROKKA (v.1.14.5) (37), any unannotated genes of interest were manually queried using BLAST. Sequence type was determined using the Achtman (38) scheme for Multi-Locus Sequence Typing (MLST, v2.0), serotype was determined by SerotypeFinder (v2.0)(39) and the average nucleotide identity (ANI) of the chromosomal genomes was calculated using the OrthoANIu algorithm (40). Plasmid replicons were determined using PlasmidFinder (v2.0.1) (41) and mobile genetic elements and resistance genes were identified using ResFinder (v4.1) (42) and Resistance Gene Identifier (RGI) (v6.0.1), using the CARD (v3.2.6) (43) database. Finally, single nucleotide polymorphisms (SNPs) and indels were identified using breseq (v0.31.1) (44).

### Nitrocefin Assay

Overnight cultures of the isolates were normalised to an optical density at 600 nm (OD600) of 0.1 in 10 ml of LB broth and centrifuged at 4,500 xg for 15 minutes. The supernatant was discarded, and the pellet resuspended in 5ml of phosphate-buffered saline (PBS). The resuspension was then sonicated on ice, for three intervals of 15 seconds, with a 30 second break between each sonication, using a Soniprep 150 plus (MSE centrifuges, UK). 90 μl of the supernatant was added to 10 μl of a 0.5 mg/ml nitrocefin solution in a 96 well clear, flat bottom plate. The absorbance of the plate was read at OD450 every 39 seconds for 9 minutes and 45 seconds, using a FLUOstar OMEGA spectrophotometer (BMG lab systems, Germany). Triplicate overnight liquid cultures were grown from the selection of three distinct colonies, each liquid culture was then assayed in triplicate.

### Aerobic bacterial growth assay

Overnight cultures of the isolates grown in LB were normalised to an OD600 of 0.1 in LB broth and 200 µl was added in triplicate to a 96 well clear, flat bottom plate. The plate was incubated for 24 hours at 37°C and the absorbance at OD600 was measured every hour using a FLUOstar OMEGA spectrophotometer.

### Determination of *bla*TEM-1 and IS*26* copy number

When comparing the bacterial isolates in the absence of TZP, isolates were grown overnight at 37°C in LB broth. Triplicate overnight liquid cultures were grown from the selection of three distinct colonies, which then underwent DNA extraction and subsequent qPCR.

To investigate gene copy number in the presence of increasing concentrations of tazobactam or piperacillin, bacterial isolates were grown in cation-adjusted Mueller Hinton broth (CA-MHB-2) containing either tazobactam fixed at 4 µg/ml and increasing concentrations of piperacillin, or piperacillin fixed at 8 µg/ml and increasing concentrations of tazobactam. Assays were then undertaken as per EUCAST guidelines (21) for AST testing. If growth was observed visually in the well, the culture underwent DNA extraction and subsequent qPCR. For each concentration of TZP three biological repeats were performed, using cultures which were grown from distinct colonies derived from a glycerol stock stored at -80° C.

Bacterial isolates were also grown over a period of five days and the gene copy number assessed at each 24-hour time point. Single colonies were grown in CA-MHB-2, CA-MHB-2 containing 4 µg/ml of tazobactam and 256 µg/ml piperacillin, and M9 liquid media (prepared as described in the supplementary methods, but without agar), for 24 hours at 37° C, 200 rpm. Following 24 hours incubation, cultures were vortexed and 1 µl transferred to fresh media. This process was repeated over a period of five days, following each 24-hour incubation, 1 ml of culture was removed, which then underwent DNA extraction and subsequent qPCR. For each isolate and media combination, three biological repeats were performed following inoculation with distinct colonies which were grown from a glycerol stock stored at -80° C.

### Quantitative PCR

Change in gene copy number of *bla*TEM-1 and IS*26* was determined using the ΔΔCT method for relative quantification (45) when compared to a single copy of the *uidA* housekeeping gene. The QuantiTect® SYBR Green PCR master mix (Qiagen, Germany) was used, following the manufacturer’s instructions. Primers used to detect *bla*TEM-1, IS*26* and *uidA* were as previously described (13, 46, 47) and were added to the reaction at a final concentration of 400 nM.

### Statistical analysis

Statistical analyses were performed in R studio with R (v4.1.1) (48) using the R Stats (v.4.1.1) and DescTools (v0.99.48) packages. Pairwise comparisons of means between two groups used t-tests and more than two groups used ANOVA, with Dunnet’s post hoc test used for pairwise group comparisons where ANOVA identified a significant between-group difference (ttest(), DunnettTest() and aov() functions) as indicated for the respective experiments in the corresponding figure legends. Graphs were produced using ggplot2 (v3.4.2)(49).Diagrams of the amplified genomic region were created using gggenes (v0.5.0) (50) and Clinker (v0.0.28) (49, 51).

## Results

### Clinical case summary

In January 2022 a woman in her 80’s was admitted to the Royal Liverpool University Hospital (RLUH) with a suspected disseminated bacterial infection. Blood tests on admission showed a raised white cell count and raised C-reactive protein (CRP) level (Figure S1) and subsequent clinical diagnoses were community acquired pneumonia, pyelonephritis and infective discitis (Figure S2).

Upon admission the patient was immediately started on a combination of antimicrobial therapy, which included TZP. The blood culture from the day of admission resulted in growth of *E. coli* resistant to amoxicillin, but susceptible to cefpodoxime, ciprofloxacin, gentamicin, meropenem and TZP. The patient remained clinically unwell and on day five, another blood culture was obtained which resulted in growth of *E. coli* with the same resistance profile except TZP, to which the *E. coli* was now resistant. The patient was then successfully treated with a sequential regimen of meropenem, ertapenem and ciprofloxacin. An extended clinical case summary can be found in the supplementary information.

### Characterisation of the isolates and identification of resistance mechanisms

We confirmed the phenotypic resistance profiles as susceptible (MIC = 1-2/4 μg/ml for TZP-S) and resistant (MIC = 256-512/4 μg/ml for TZP-R). The growth curves (supplementary methods) of the two isolates under standard conditions showed no significant difference (Figure S3).

Assembling the short- and long-read sequencing data resulted in a total genome length of 5,305,542 bp for the TZP-S isolate (GenBank accession ID: JAYMZA000000000). This included one conjugative plasmid 162,475 bp in length which was assembled as a single contig and contained IncFIA, IncFIB and IncFII replicons. The assembly of the TZP-R isolate (GenBank accession ID: JAYMZB000000000) resulted in a total genome length of 5,353,725 bp and included a larger conjugative plasmid (assembled as one contig) of 210,651 bp and which contained the same replicons as the TZP-S isolate. Also assembled within the TZP-R genome was an additional smaller circular DNA molecule of 54,316 bp; however due to lack of sequencing depth of both long and short reads, we could not confidently ascertain the structure of this region. A plasmid replicon was not detected within this molecule, and it was not predicted to contain any antimicrobial resistance genes (ARGs) though it was predicted to contain a single copy of *ompX*, an outer membrane porin along with multiple copies of IS*21*, an exonuclease, and phage-related assembly and integrase genes.

We determined the isolates to belong to sequence type 131 and serotype O153:H4. We also confirmed the clonality of the TZP-S and TZP-R isolates, as the ANI of the chromosomal genomes was calculated as 99.99% and only three SNPs were detected between them on the chromosome. Both were predicted to contain the β-lactamase *bla*TEM-1; however, whilst the TZP-S isolate was predicted to contain one copy of the gene (Figure 1A), the TZP-R isolate was predicted to contain ten copies (Figure 1C). In both cases *bla*TEM-1 was located on the large conjugative plasmid, as part of a resistance cassette (Figure 1B) that contained a transposase gene, along with *dosP*, *merE, merD* and *lpd1.* In addition, a hypothetical protein, which when manually queried was described as a “mobile element protein” and *pinE*, which encodes a site-specific DNA recombinase, were also annotated only once in the resistance cassette of both the TZP-S and TZP-R isolate. These were not duplicated. In both the TZP-S and TZP-R isolate the resistance cassette contained flanking copies of IS*26,* which were oriented in opposing directions. The IS*26* elements located within the duplicated region in the TZP-R isolate were oriented in the same direction. The *bla*TEM-1 genes in both isolates are under control of the strong P4 promoter (52) and no SNPs were detected within the promoter region or elsewhere within the plasmid.

**Figure 1.**
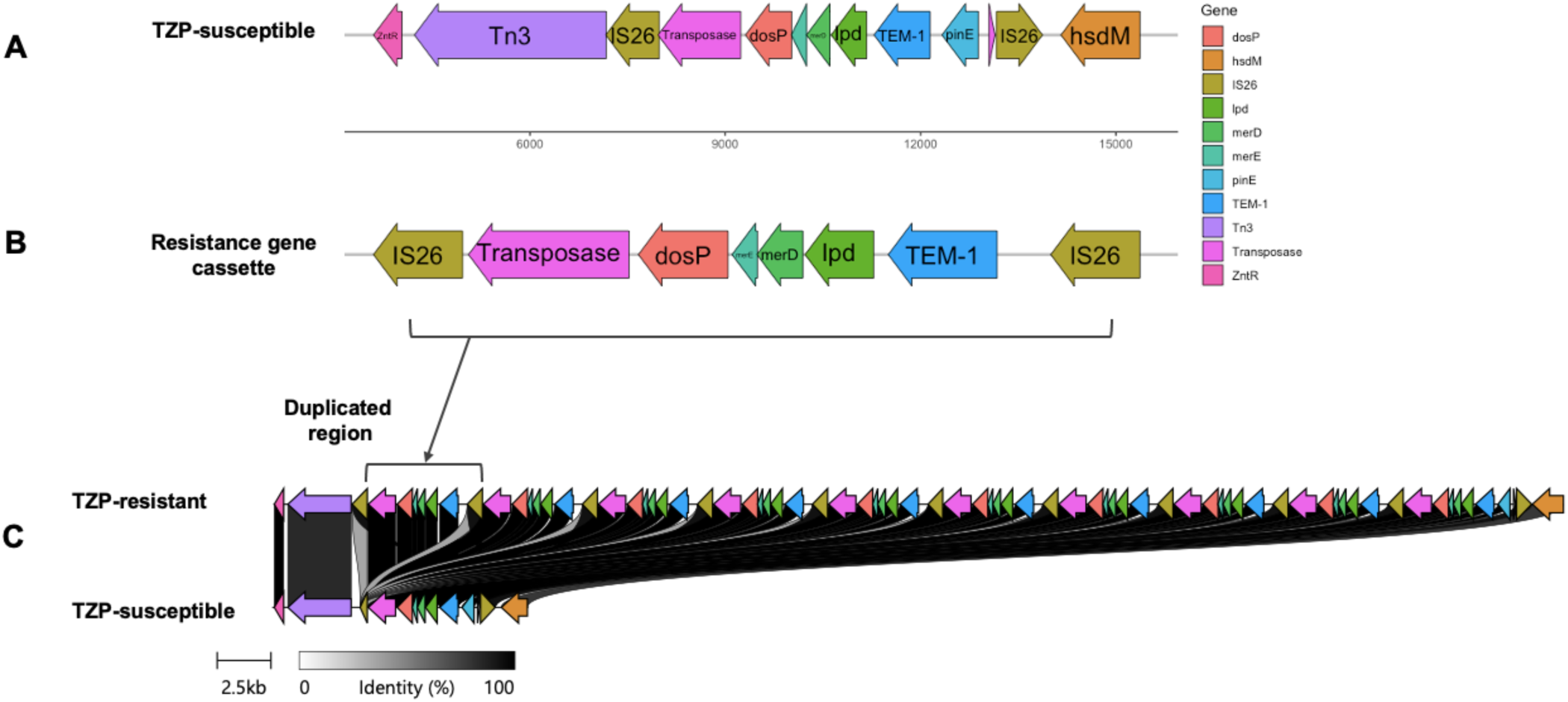
Schematic showing **(A),** the location of resistance cassette within the TZP-S isolate **(B)** the structure of the resistance gene cassette which was duplicated ten times in the TZP-R isolate, and **(C)** the size and percentage identity of the duplicated region in the TZP-R isolate, when compared to the TZP-S isolate.

Comparison of the predicted resistance genes in both isolates highlighted the presence of resistances to additional drug classes (Table 1) located both on the plasmid and the chromosome. Other than duplication of *bla*TEM-1, no differences in ARGs, including resistance-conferring SNPs, were predicted when comparing the TZP-S isolate with the TZP-R isolate.

**Table 1.** The TZP-S and TZP-R isolates had the same predicted repertoire of ARGs, the only exception was the number of copies of *bla*TEM-1, one copy was found in the TZP-S isolate, whereas ten copies were found in the TZP-R isolate. RGI was used to identify genes, using the CARD database, only genes designated as perfect hits (100% sequence identity and coverage) are shown.

| Resistance genes/mutations | Location | Drug class |
| --- | --- | --- |
| *bla*<sub>TEM-1</sub> | Translocatable unit | β-lactam |
| *aadA5* | Plasmid | Aminoglycoside |
| *qacEdelta1* | Plasmid | Antiseptics/disinfectants |
| *sul1* | Plasmid | Sulfonamide |
| *Mrx* | Plasmid | Macrolide |
| *mphA* | Plasmid | Macrolide |
| *evgA* | Chromosome | Macrolide, fluoroquinolone, penam, tetracycline |
| *baeR* | Chromosome | Aminoglycoside, aminocoumarin |
| *H-NS* | Chromosome | Macrolide, fluoroquinolone, β-lactam, tetracycline |
| *mdtH* | Chromosome | Fluoroquinolone |
| *leuO* | Chromosome | Nucleoside, antiseptics/disinfectants |
| *gadW* | Chromosome | Macrolide, fluoroquinolone, penam, tetracycline |
| *cpxA* | Chromosome | Aminoglycoside, aminocoumarin |

### Confirmation of increased β-lactamase activity

Having identified the duplication of *bla*TEM-1 in the TZP-R isolate, we used the nitrocefin hydrolysis assay to determine the comparative activity of β-lactamase. Following overnight culture of both isolates in nutrient rich media and in the absence of TZP a significant increase in nitrocefin hydrolysis was observed in the TZP-R isolate at the midway point of the assay (P < 0.001), confirming increased levels of β-lactamase enzymatic activity (Figure 2A and 2B). qPCR performed on the isolates showed increased gene copy number. The log2 fold change of both *bla*TEM-1 and IS*26* (relative to the single-copy housekeeping gene *uidA*) were significantly increased from 0.97 to 7.05 (p = 0.001, log2 fold change = -0.05 to 2.80) and 3.16 to 8.20 (p = 0.003, log2 fold change = 1.66 to 2.89) respectively, in the TZP-R isolate (Figure 2C).

**Figure 2.**
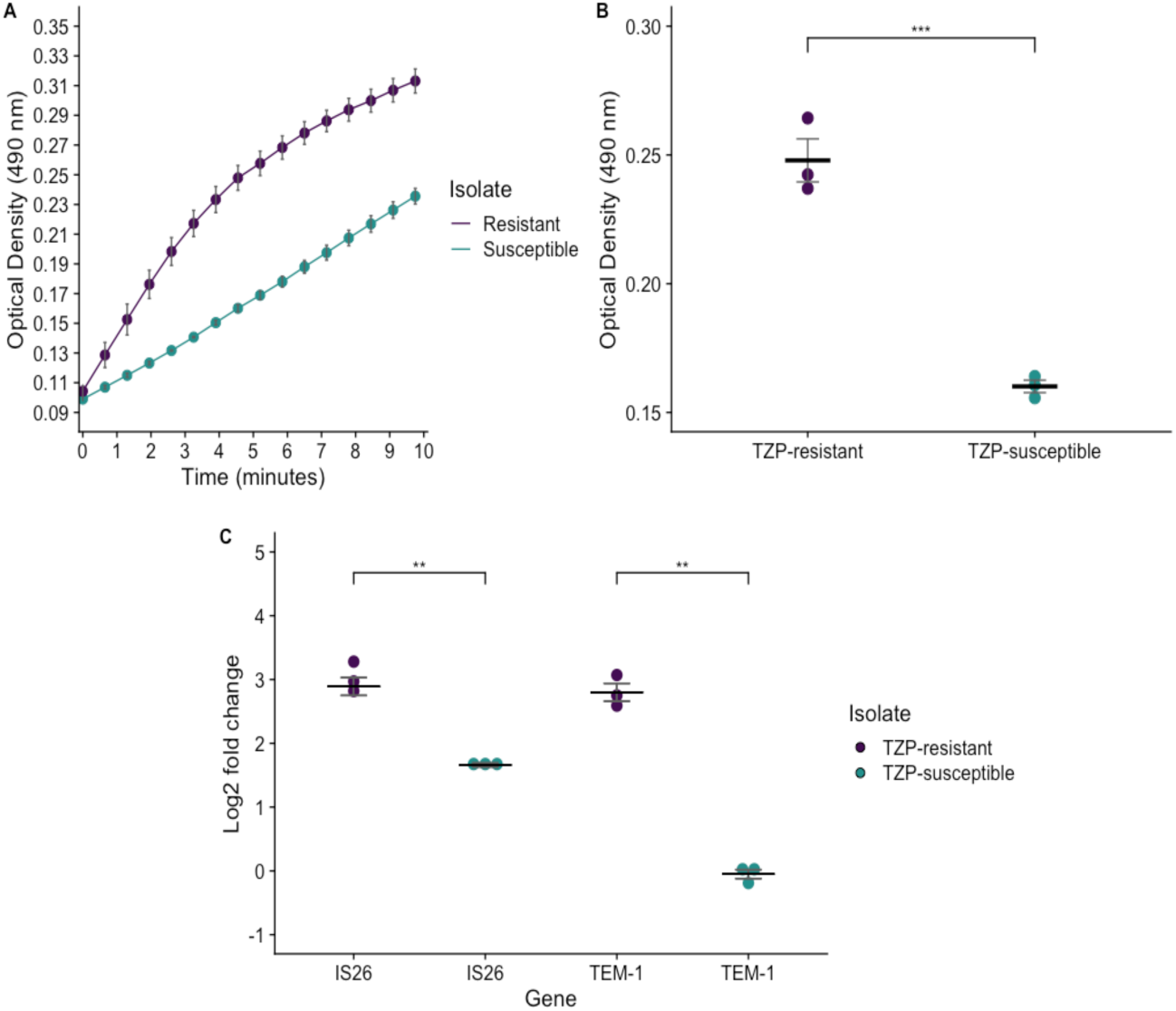
Nitrocefin hydrolysis measured at OD450 was greatest in the TZP-R isolate (purple points/line) throughout the assay **(A),** and significantly increased at the middle time-point of the assay **(B),** when compared to the TZP-S isolate (green points/line). qPCR performed showed an increase in gene copy number of both *bla*TEM-1 and IS*26* in the TZP-R isolate, when compared to the TZP-S isolate, following normalisation against *uidA,* a single-copy housekeeping gene (**C)**. In both experiments, three biological repeats using distinct bacterial colonies were performed and each measurement was made up of three technical repeats. In **(A)** points show the mean of the three biological repeats, which had been determined from the technical repeats. In **(B)** and **(C)** each point shows an individual biological repeat, the black horizontal like shows the mean of the biological repeats. In all cases error bars show the standard error of the mean (SEM). Statistical significance in **(B)** and **(C)** was calculated using an unpaired t-test. For **(C)** non-transformed copy number estimates were used to determine statistical significance ** = P ≤ 0.01, *** = P ≤ 0.001.

### Copy number of *bla*TEM-1 and IS*26* in response to an increasing concentration of tazobactam

We aimed to determine if increasing the concentration of tazobactam, whilst fixing the piperacillin at the breakpoint (8 μg/ml), led to a change in *bla*TEM-1 and IS*26* copy number. Copy number of *bla*TEM-1 in the TZP-S isolate increased when the concentration of tazobactam was 1 μg/mL or greater (Figure 3). However, a significant increase in copy number was only observed when analysing the gene copy number change in response to 2 μg/ml, when compared to 0 μg/ml of tazobactam. Fold change of *bla*TEM-1 compared to the single-copy housekeeping gene *uidA* increased from 0.82 to 1.44 (p = 0.0065, log2 fold change = -0.31 to 2.63). Copy number of IS*26* remained similar in the TZP-S isolate between tazobactam concentrations of 0 to 1 μg/ml, however a statistically significant increase in copy number of IS*26* was observed with 2 μg/ml tazobactam where fold change increased from 2.51 to 4.66 (p = 0.0067, log2 fold change 1.33 to 2.22). Similarly, in the TZP-R isolate, copy number of *bla*TEM-1 increased when the concentration of tazobactam was 1 μg/mL or greater (Figure 3) and a statistically significant increase in copy number was only observed when analysing the change in response to 8 μg/ml and compared with 0 μg/ml of tazobactam. Fold change of *bla*TEM-1, compared to the single-copy housekeeping gene *uidA,* increased from 6.24 to 10.95 (p = 0.001, log2 fold change = 2.62 to 3.45). In the TZP-R isolate a similar copy number of IS*26* was observed across concentrations of tazobactam from 0-8 μg/ml and no significant changes in *IS26 copy* number were identified.

**Figure 3.**
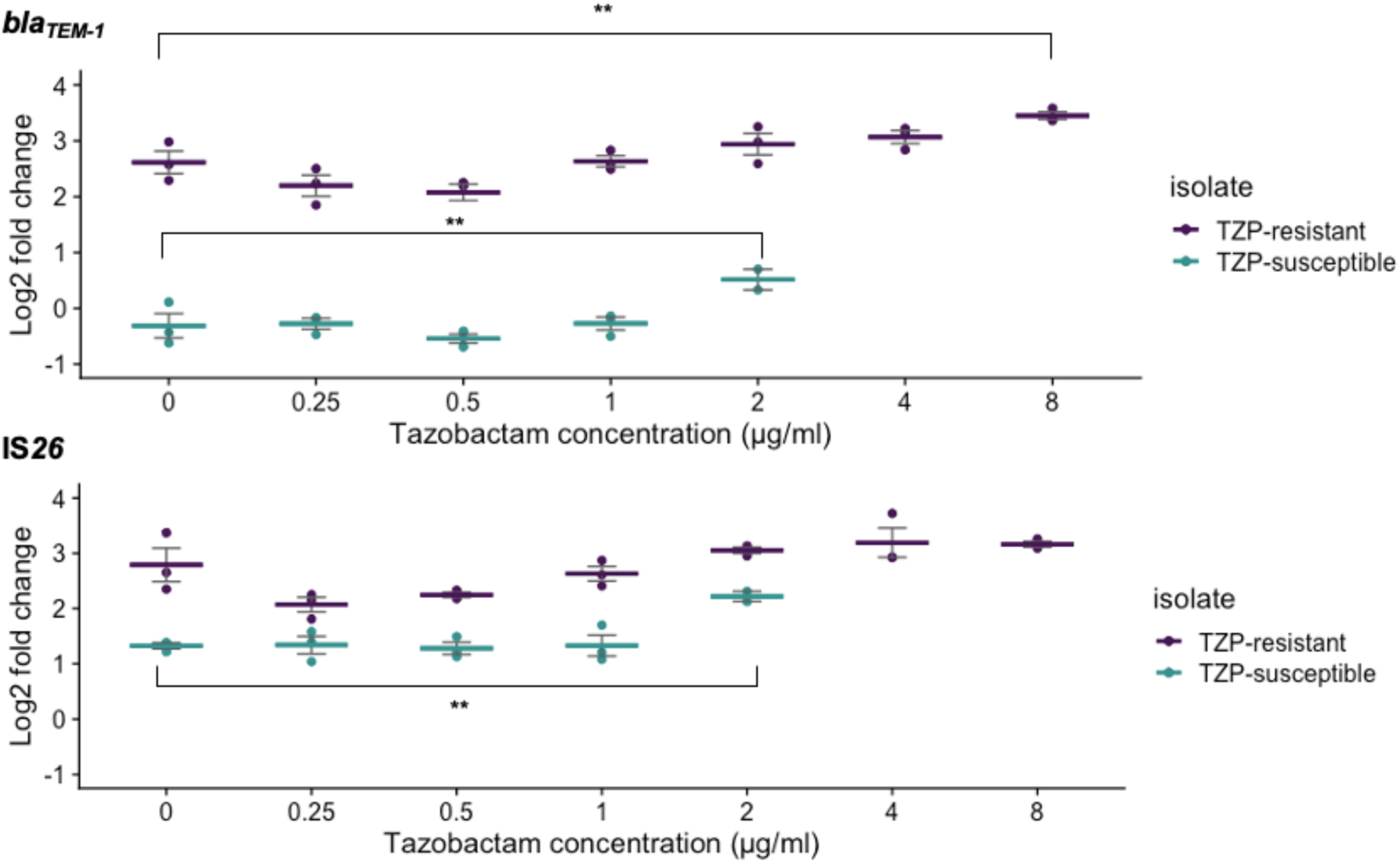
qPCR performed following the culture of isolates in an increasing concentration of tazobactam showed that gene copy number of *bla*TEM-1 in both the TZP-S (green points/dashes) and TZP-R (purple points/dashes) isolate was greatest when the concentration of tazobactam was also greatest. A significant increase in IS*26* copy number was only detected in the TZP-S (green points/dashes) isolate when the tazobactam concentration was 2 μg/ml. The TZP-S isolate was not able to survive at concentrations of tazobactam which exceeded 2 μg/ml. The concentration of piperacillin was fixed at 8 μg/ml and *bla*TEM-1 and IS*26* copy number and fold change were calculated following normalisation against *uidA,* a single-copy housekeeping gene. Each point shows one biological repeat, and each dash shows the mean of the three biological repeats. Error bars show the SEM. Graphs showing each biological repeat in isolation can be found in the supplementary, (Figure S4). Statistical analysis was performed on non-transformed copy number estimates using ANOVA and Dunnett’s post hoc test using 0 μg/ml of piperacillin as the control group. *** = P ≤ 0.01.

### Copy number of *bla*TEM-1 and IS*26* in response to an increasing concentration of piperacillin

As the TZP-R isolate survived in concentrations of piperacillin up to and greater than 256 μg/ml, we investigated the relationship between increasing piperacillin concentration and the copy number of *bla*TEM-1 and IS*26* whilst fixing the concentration of the inhibitor. Overall, copy numbers of both *bla*TEM-1 and IS*26* did increase in response to an increasing concentration of piperacillin (Figure 4). However, a statistically significant increase in gene copy number was only observed when analysing the gene copy number change in response to 256 μg/ml, when compared with 0 μg/ml of piperacillin. When comparing these conditions, fold change of *bla*TEM-1 and IS*26* compared to the single-copy housekeeping gene *uidA* increased from 9.47 to 53.95 (p = <0.01, log2 fold change = 3.22 to 5.58), and 8.83 to 38.21 (p < 0.01, log2 fold change = 3.12 to 5.17), respectively. This indicates a step-change of gene copy number rather than a gradual increase, in response to increasing piperacillin concentration.

**Figure 4.**
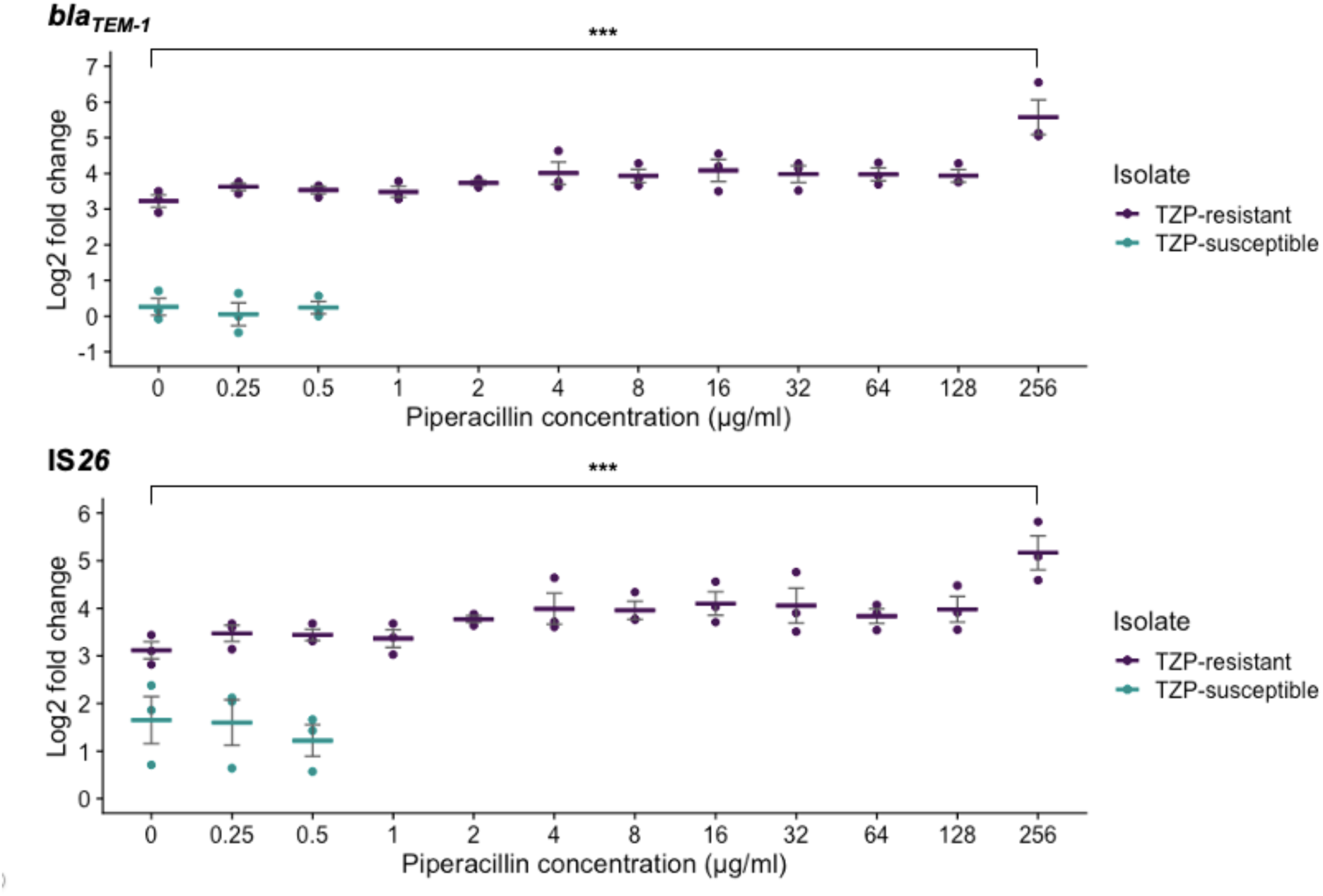
qPCR performed following the culture of isolates in an increasing concentration of piperacillin, showed that gene copy number both *bla*TEM-1 and IS*26* in the TZP-R (purple points/dashes) isolate was greatest when the concentration of piperacillin was also greatest. The TZP-S isolate (green points/dashes) was not able to survive at concentrations of piperacillin which exceeded 0.5 μg/ml. However, when the concentration of piperacillin was 0, 0.25 or 0.5 μg/ml, a significant increase in copy number of *bla*TEM-1 and IS*26* was detected in the TZP-R isolate. The concentration of tazobactam was fixed at 4 μg/ml and *bla*TEM-1 and IS*26* copy number and fold change were calculated following normalisation against *uidA,* a single-copy housekeeping gene. Each point shows one biological repeat, and each dash shows the mean of the three biological repeats. Error bars show the SEM. Graphs showing each biological repeat in isolation can be found in the supplementary, (Figure S5). Statistical analysis was performed on non-transformed copy number estimates using ANOVA and Dunnett’s post hoc test using 0 μg/ml of piperacillin as the control group. *** =

### Copy number of *bla*TEM-1 and *IS26* remains stable in the TZP-R isolate for a period of five days in the absence of TZP

To investigate the stability of the amplification of *bla*TEM-1 and IS*26* and whether it was sustained over multiple daily passages, we determined their copy number over five days with daily sampling and qPCR. This was measured both in the presence or absence of selection pressure (CA-MHB2 supplemented with TZP or without, respectively) and a nutrient-limited (MM) environment (Figure 5). Gene copy number of both *bla*TEM-1 and IS*26* was highest in the TZP-R isolate when cultured in CA-MHB2 supplemented with TZP. In the presence of TZP there was an increase in gene copy number of *bla*TEM-1 and IS*26* from day one to day two, which was sustained until day five (Table 2). In the absence of TZP gene copy number remained stable, with no significant change in either *bla*TEM-1 or IS*26,* at any day during the experiment, compared to the day one. This indicates that removal of selection does not readily reverse this amplification and the resistance phenotype. This is also consistent with the lack of a measurable growth fitness cost of TZP-R compared to TZP-S.

**Figure 5.**
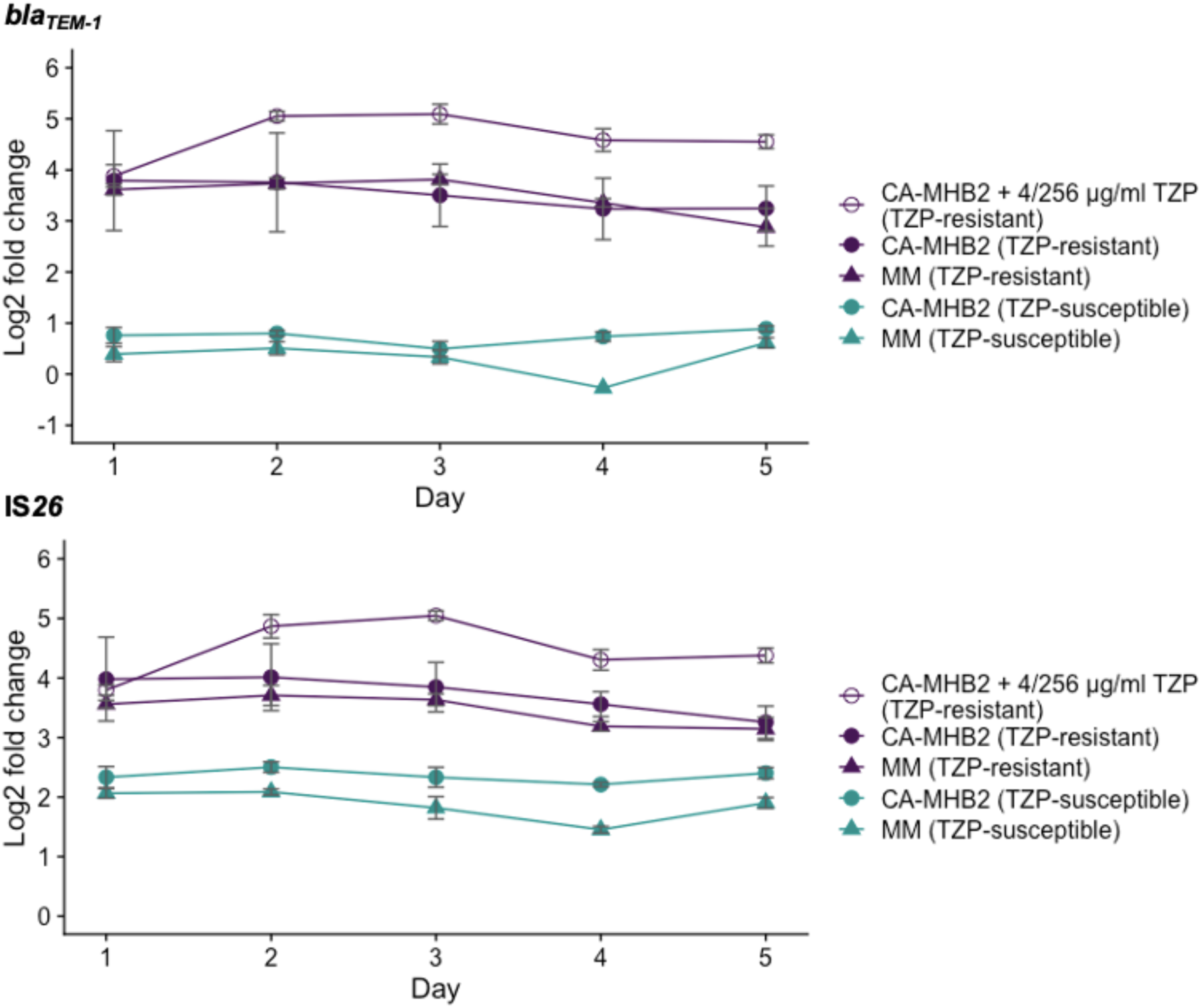
qPCR performed following the passage of isolates every 24 hours, over a period of five days showed that gene copy number both *bla*TEM-1 and IS*26* in the TZP-R (purple) isolate was greatest when MHB was supplemented with TZP (open purple circles). In both the TZP-R isolate and the TZP-S (green) isolate copy number of both *bla*TEM-1 and IS*26* remained stable over the experimental period. Copy number and fold changeof *bla*TEM-1 and IS*26* were calculated following normalisation against *uidA,* a single-copy gene. Each point shows the mean of three biological repeats. Error bars show the SEM.

**Table 2.** Gene copy number increase of *bla*TEM-1 and IS*26* in the presence of 4/256 μg/ml TZP. The TZP-R isolate was passaged in fresh MHB + 4/256 μg/ml TZP every 24 hours, for a period of five days. Copy number was estimated from qPCR and calculated following normalisation against *uidA,* a single copy gene. Statistical analysis was performed on non-transformed gene copy number estimates using ANOVA and Dunnett’s post hoc test, using day one as the control group.

| **A** *bla*<sub>TEM-1</sub> |  |  | **B** *IS26* |  |  |
| --- | --- | --- | --- | --- | --- |
| Day | Gene copy number increase | P value | Day | Gene copy number increase | P value |
| 1-2 | 18.26 | < 0.01 | 1-2 | 15.80 | <0.01 |
| 1-3 | 19.66 | < 0.01 | 1-3 | 19.20 | <0.001 |
| 1-4 | 9.46 | NS | 1-4 | 6.09 | NS |
| 1-5 | 8.60 | NS | 1-5 | 6.98 | NS |

### Investigation into other genes duplicated within the resistance cassette

We next explored the effect of the duplication of other genes in the TZP-R duplicated transposable unit, in particular *dosP* and *lpd*. The co-amplification of these genes with ARGs, facilitated by MGEs, has not previously been described. DosP is a well characterised phosphodiesterase, which is usually encoded as part of the *dosCP* operon and regulates cellular levels of the secondary messenger, c-di-GMP by hydrolysing it to linear-di-GMP (53, 54), which is implicated in controlling biofilm formation and swimming motility (55). However, though *dosP* was duplicated ten-fold in the TZP-R isolate, we found no significant difference in either motility (supplementary methods, Figure S6A) or biofilm production (supplementary methods, Figure S6B) when comparing the two isolates, and no change in the *dosCP* operon on the chromosome.

The Congo red assay (supplementary methods), which differentially stains extracellular matrix components in live cells, highlighted a visual difference in morphology when comparing the two isolates (Figure S7). Following 24 hours growth, the TZP-S isolate had bound very little of the dye and colonies appeared pale white/cream in colour (Figure 7A). In contrast, the TZP-R isolate had taken up the dye and colonies were predominantly dark-red in colour and had a distinctive star like pattern, with lines radiating out from the centre of the colonies, which previous studies have indicated occur due to cellulose production (56, 57). As *dosP* decreases c-di-GMP levels, and c-di-GMP stimulates cellulose synthesis, increased expression of *dosP* should reduce cellulose production (54, 58, 59), thus contradicting the morphotype we observed. Upon reviewing the SNP analysis, we did not find supporting evidence of SNPs which may alter the production of cellulose and could not find a genomic basis for the morphotype.

Along with *dosP*, *lpd*, which encodes dihydrolipoamide dehydrogenase (LPD), was also duplicated ten-fold. LPD is critically important for the regulation of *E. coli* metabolism in anaerobic conditions (60–62), so we assessed the growth of the two isolates under anaerobic conditions. However, we found no significant difference in growth, following incubation on either M9 agar, or MHA, in anaerobic conditions (Figure S8).

Following previous unsuccessful attempts to identify a phenotype change linked to the amplification of *dosP* and *lpd*, we conducted a broad screening to assess the isolates’ ability to utilize different carbon and nitrogen sources. No significant differences were observed between the TZP-S and TZP-R isolates in the carbon utilization assay (PM1) (Figure S9).

Similarly, no meaningful differences were found between the TZP-S and TZP-R isolates in the nitrogen utilization assay (PM3b) (Figure S10). However, due to the variance observed between biological replicates (Figures S12 and S14) for some of the tested compounds, we were unable to confidently establish average growth curves in response to those compounds.

## Discussion

We performed comparative genome sequence and phenotype analyses on two patient-derived isolates that underwent within-patient evolution during antimicrobial treatment. We confirmed that the greater than 100-fold difference in MIC to TZP between the isolates at the beginning and after several days of antimicrobial treatment, respectively, is due to *bla*TEM-1 duplication and subsequent enzyme hyperproduction. We observed that ten copies of *bla*TEM-1 are sufficient to induce this high level of enzyme production and resistance to TZP, but that susceptibility to 3GCs and carbapenems is retained. We also found that culture of the TZP-R bacteria in high levels of piperacillin or tazobactam both further increased the copy number of *blaTEM-1*. Like other studies (13–15), we found that the extensive amplification of *bla*TEM-1 was mediated by IS*26* elements. The transmissible nature of IS*26* plays a key role in the dissemination of AMR genes (63), and the location of the resistance cassette on a plasmid also highlights the potential for both intraspecies and interspecies movement of *bla*TEM-1.

Recent reports have characterised the amplification of *bla*SHV-5 (64), inducing resistance to 3GCS, and *bla*CMY-146 (65) and *bla*CTX-M-15, inducing resistance to carbapenems(66)

There are conflicting data which describe fitness cost associated with TEM-1 hyperproduction when induced by gene duplication (13, 14). We found that gene copy number of *bla*TEM-1 and IS*26* was stable over a period of five days, even in the absence of selection pressure. In this study the TZP-R isolate contained additional 48,195 bp within the plasmid compared to the TZP-S isolate, including multiple additional coding sequences. However, when comparing the growth dynamics of the two isolates we did not find a metabolic cost associated with replicating an enlarged plasmid, as the growth curves of the two isolates were comparable; however, we acknowledge that this is a simplistic model which is unlikely to replicate the environment within a human host. As we only confirmed the increased expression of *bla*TEM-1 and IS*26* it is possible that the additional duplicated genes located on the resistance cassette were not expressed at high levels, however it is also plausible that the metabolism-linked *lpd* gene duplicated via IS*26* as part of the *bla*TEM-1 cassette provided a compensatory fitness advantage to the TZP-R isolate, but detailed investigation of this goes beyond this study.

Another gene upregulated as part of the IS26-induced *bla*TEM-1 cassette amplification was *DosP*. We note that *DosP* has a role in c-di-GMP metabolism, a complex key regulator of many other cellular processes including growth (53), stress adaptation (67), persistence (68) and virulence (55); in particular the observation of increased persistence of *E. coli* in ampicillin and ciprofloxacin following overexpression of *DosP* (61), which could be driven by the extensive duplications of *dosP* we observed in the resistant isolate. We could not find any significant difference in motility or biofilm production, two processes regulated by DosC and DosP, which might be due to the low numbers of *DosC* compared to *DosP* in the resistant isolate. As we did not quantify the ratio of c-di-GMP to linear-di-GMP we cannot be sure that the extensive duplication of *dosP* resulted in increased c-di-GMP hydrolysis.

The morphology of the TZP-R isolate on congo-red agar was markedly different from the TZP-S isolate; and whilst the colonies did not appear “rough” on visualisation, there are marked similarities with the “pink/red, dry and rough” (rdar and pdar) morphotype observed in other *Enterobacterales.* Rdar morphology has been reported in *Salmonella* (*69*)*, E. coli* (*55, 56*) and *Klebsiella pneumoniae* (70) and is associated with increased cellulose expression, which can improve bacterial persistence in adverse environmental conditions, and is understood as a step in adaptation to the human host (71). Confirmatory experiments are needed, however we hypothesize that the morphology of the TZP-R isolate may be due to increased cellulose expression, though we are currently unable to find a genomic explanation for this. Our study further emphasizes the relevance to better understand and try to prevent resistance phenotype evolution as a result of antimicrobial treatment, as the evolutionary pressure can lead to the emergence of not only more resistant, but also better host-adapted and putatively more virulent strains.

A broader assessment of carbon and nitrogen utilization yielded no meaningful findings. However, we acknowledge that the variance among replicates for the nitrogen utilization plates may have obscured subtle differences. Additionally, we did not subject the isolates to any antimicrobial selection pressure, which might have been necessary to trigger the pathways responsible for inducing a modified phenotype.

The importance of improving phenotypic-based AST, biochemical assays and utilising predictions from genomic data is highlighted by the TZP-R/3GC-S phenotype presented in this study, and the need to account for gene copy number and predicted expression levels when predicting resistance to BL/BLI combinations. Scores for BL/BLI resistance prediction from genomic data have been developed (72) in addition to β-lactamase copy number calculators (73) and tools like these will be vital to support decisions for use of new BL/BLI combinations and ensure their future efficacy is safeguarded. Alternative diagnostic methods, based on protein detection, such as matrix-assisted laser desorption/ionization mass spectrometry (MALDI-MS) technology, may provide an alternative strategy to rapidly detect β-lactamase hyperproduction (74).

BL/BLI combinations such as TZP are an important strategy for the treatment of invasive and disseminated infections caused by *E. coli*, including those caused by *E. coli* strains expressing Ambler class A β-lactamases such as *bla*TEM-1 (5). Our investigation contributes to a growing body of evidence highlighting hyperproduction of TEM-1 as a mechanism by which tazobactam use can be overcome, resulting in treatment failure. In the absence of other information, TZP treatment failure may incorrectly suggest that an isolate is an ESBL- or ampC producing strain and promote carbapenem usage. In this study, a carbapenem was ultimately used and treatment was successful, however it may have also been possible to successfully treat the patient with a 3GC. Promoting the de-escalation of antimicrobial selection and reducing exposure to broad-spectrum antimicrobials of last resort (such as carbapenems) is vitally important to prevent the further emergence of resistance and retain viable treatment options. It is therefore imperative that alternative mechanisms of AMR, such as IS-mediated β-lactamase amplification are a priority area of research, especially efforts focused of the identification of these genotypes and understanding their evolution and transmission.

## Data Availability

All data produced in the present study are available upon reasonable request to the authors and WGS can be found at NCBI under the bioproject ID PRJNA1061590.

## Acknowledgments

We would like to thank Dr Alan Cartmell for generously providing the *Bacteroides thetaiotaomicron* (VPI-5482) positive anaerobic control strain. We would like to thank the patient for their collaboration and enabling our research.

