## Supplementary data and methods for "A high-resolution genomic and phenotypic analysis of resistance evolution of an *Escherichia coli* strain from a critical care patient treated with piperacillin/tazobactam"

### Supplementary information

#### Supplementary methods

##### Biofilm assay

Colonies grown overnight in LB broth were normalised to an $OD_{600}$ of 0.1 in both M9 media and cation adjusted-Mueller Hinton broth (CA-MHB) and 150 µl of inoculated media was then transferred to a non-treated, 96-well U-bottomed clear plate. Plates were then incubated at 37°C for 24 hours. Wells were then washed three times with PBS, stained with 0.1% crystal violet (Sigma, U.K.) and left to incubate for 20 minutes. Wells were then washed a further three times with PBS, before the strain was solubilised using 30% acetic acid (Sigma, U.K.). Following a 15 minute incubation, 125 µL of liquid from the wells was transferred to a flat-bottomed clear plate and the absorbance read at $570_{nm}$ using a SPECTROstar OMEGA spectrophotometer.

##### Motility assay

Swimming motility agar plates contained 25 ml of media, comprised of 1% tryptone, 0.5% yeast extract, 0.5% NaCl and 0.3% agar (all Sigma, U.K.). Overnight cultures were normalised to an $OD_{600}$ of 0.1 in PBS and 2 µl of culture was inoculated into the agar in the centre of the plate. Plates were then incubated at 37°C for 24 hours. The plates were then photographed, and the area calculated using imageJ software (v153), using the diameter of the plate as a scale reference. For each isolate, three biological repeats consisting of three technical repeats were performed.

##### Congo red plate assay

LB agar was supplemented with Brilliant Blue G-250 and Congo Red, to a final concentration of 20 µg/ml and 40 µg/ml, respectively. Overnight cultures grown in LB were diluted to $10^6$ in PBS and 50 µl was transferred to the plates which were then incubated for a total of 48 hours at 37°C. The colonies were observed using a GXMXTL101LED Stereozoom microscope and photographed at 24 hours and 48 hours using a GXCAM 5MP USB-3, C-Mount Microscope camera. For each isolate, three biological repeats, consisting of three technical replicates were performed.

##### Anaerobic bacterial growth assay

Colonies grown on LB agar were suspended in PBS and normalised to an $OD_{600}$ of 0.1. Sterilised nitrocellulose filters (0.45 µM pore-size) (Cytiva, USA), were placed on plates containing Mueller Hinton Agar (MHA) or M9 agar (20% (v/v) M9 minimal salts (5x), 0.4% D-glucose, 4 mM magnesium sulphate, 0.05 mM calcium chloride and 1.5% agar) (all Sigma,

U.K.) and 10 µl of the bacterial suspension was spotted on top of the filter. Plates were then incubated at 37° for 24 hours, either in the presence of oxygen; or in an anaerobic environment produced by using the Oxoid™ Anaerojar ™system (Thermo Scientific, U.K.). Following incubation, the filters were removed from the agar media and placed in 10 ml PBS which was then thoroughly vortexed. Fifty µl of serially diluted ($10^{-4}$, $10^{-5}$ and $10^{-6}$ ) resuspended bacteria was then plated on LB agar, the plates were then incubated aerobically at 37°C for 24 hours. Following incubation, colonies were counted and related back to CFU/ml. The experiment was performed on three separate occasions, each time three technical repeats were performed. Alongside the biochemical anaerobic indicator, the obligate anaerobe *B. thetaiotaomicron* was grown to confirm the generation of an anaerobic environment.

**Biolog assay for carbon and nitrogen utilisation**

Isolates were grown on LB overnight at 37°C. Inoculations to Biolog phenotypic microarray plates were set up following manufacturer's instructions with below modifications. Biomass was suspended to an O.D.600 of 0.040 +/- 0.005 in complete inoculation fluid as indicated in table S1. Metal ion cocktail contained 5 mM each of $ZnCl_2$ $7H_2O$, FeCl2 $6H_2O$, $MnCl_2$ $4H_2O$, $CaCl_2$ $2H_2O$ and were filter sterilized using a 0.22 µM polyethersulfone filter. To wells of each PM1 or PM3b plate (Techno-path, 12111 or 12121, respectively), 100 µl of this suspension was added and strains incubated for up to 72 hours at 37°C, static, in a OmniLog PM system (imaging every 15 minutes). Data was extracted using Biolog softwares (conversion of D5E to OKA: D5E_OKA Data File Converter v1.1.1.15 and extraction of raw kinetic data using PM analysis software: Kinetic V1.3). Plots were produced using ggplot2 (v3.4.2) using R (version 4.3.2). Useful experimental metrics were acquired using the BactEXTRACT webtool (https://www.microbiologyresearch.org/content/journal/acmi/10.1099/acmi.0.000742.v1)
.

**Table S1**

| Ingredient | PM1 | PM3 |
|---|---|---|
| IF-0a, Techno-path, 72268 | 10 mL | 10 mL |
| Dye Mix D, Techno-path, 74221 | 0.12 mL | 0.12 mL |
| Sodium succinate (20 mM | - | 0.12 mL |
| Metal ion cocktail (50 µM)* | 0.12 mL | 0.12 mL |
| Water | 0.60 mL | 0.60 mL |
| Cell suspension | 1.16 mL | 1.16 mL |
| Total | 12 mL | 12 mL |

**Supplementary information and figures.**

**Extended clinical case summary**

Clinical assessment and investigations undertaken upon the patient's presentation at the RLUH, including a chest X-ray, led to an initial diagnosis of community acquired pneumonia (CAP). Further investigations indicated a disseminated infection, as a computed tomography (CT) scan was consistent with pyelonephritis in the right kidney and a magnetic resonance imaging scan (MRI) showed infective discitis (supplementary figure 2). Blood tests on admission showed a raised white cell count and raised C-reactive protein (CRP) level, indicating likely infection (Figure 1). The white cell count (figure 1B) fluctuated between clinically elevated and clinically normal levels throughout the 23 days that followed admission; CRP levels (figure 1C) decreased over the same period, though remained slightly elevated at day 23 post admission.

Samples for blood cultures were taken on initial examination on day one (figure 1B/C) of admission and intravenous (IV) TZP was prescribed immediately, which was continued until day seven. Due to suspected CAP, amoxicillin, clarithromycin and gentamicin were also given intravenously. The blood culture from the day of admission resulted in growth of *E. coli* resistant to amoxicillin, but susceptible to cefpodoxime, ciprofloxacin, gentamicin, meropenem and TZP. As the patient continued to have intermittent pyrexia of >38°C and was still clinically unwell, further doses of IV gentamicin were given on day three and five of admission. On day five, another blood culture (figure 1B/C) was obtained which resulted in growth of *E. coli* with the same resistance profile except TZP, to which the later isolate was resistant. Antimicrobials were changed to meropenem on day seven of admission and were given for three days, before changing to oral ciprofloxacin for three days. In response to spinal tenderness, oedema and signs of infection indicated on the MRI of the spine, antimicrobials were changed back to IV meropenem, until day 22 when they were changed to ertapenem, the patient was then given oral ciprofloxacin for a further four weeks. The patient was reviewed a month after ending antimicrobial therapy, CRP levels were normal and there was no evidence of persistent infection.

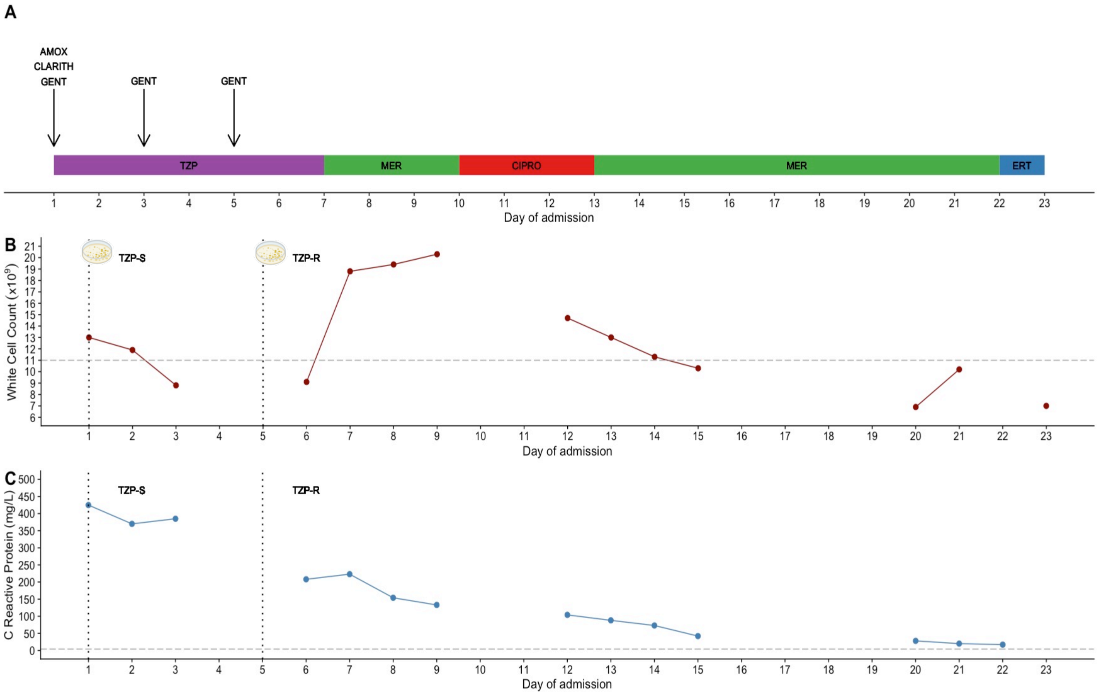


**Figure S1.** Clinical laboratory results and antimicrobial prescribing data for the patient from whom the *E. coli* isolates were obtained, from day 0 to day 22 of admission. Antimicrobials prescribed **(A)** included piperacillin/tazobactam (TZP), amoxicillin (AMOX), clarithromycin (CLARITH), gentamicin (GENT), ciprofloxacin (CIPRO), meropenem (MERO) and ertapenem (ERT). White cell count **(B)** was deemed elevated if it was above $11x10^9$/L, which is indicated by the horizontal grey dashed line. C reactive protein **(C)** was deemed elevated if it was above 4 mg/L, which is indicated by the horizontal grey dashed line. The days on which blood cultures were obtained, which led to the classification of the *E. coli* isolate as either TZP-susceptible (TZP-S), or TZP-resistant (TZP-R) is indicated on **(A)** and **(B)** by the dashed vertical black lines.

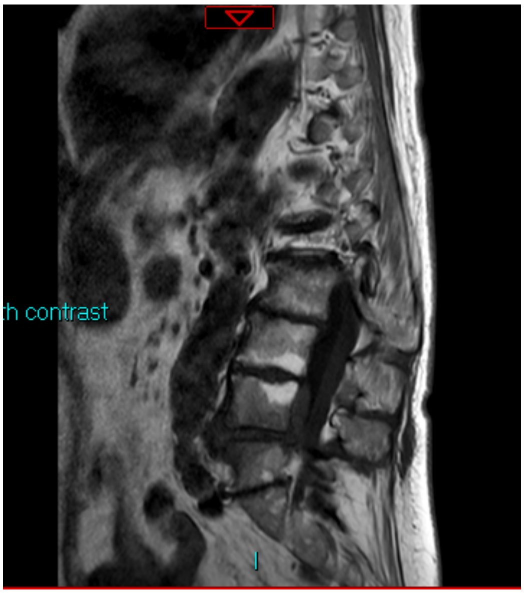


**Figure S2.** An MRI spine was performed on day 12 due to lumbar spine tenderness which demonstrated oedema of the L4/5 intervertebral disc and adjacent endplates in keeping with infective discitis, an extradural collection measuring 7 mm (AP) x 48 mm (CC), posterior to the L4 and L5 vertebral bodies and at least moderate spinal canal stenosis at L4/5 level.

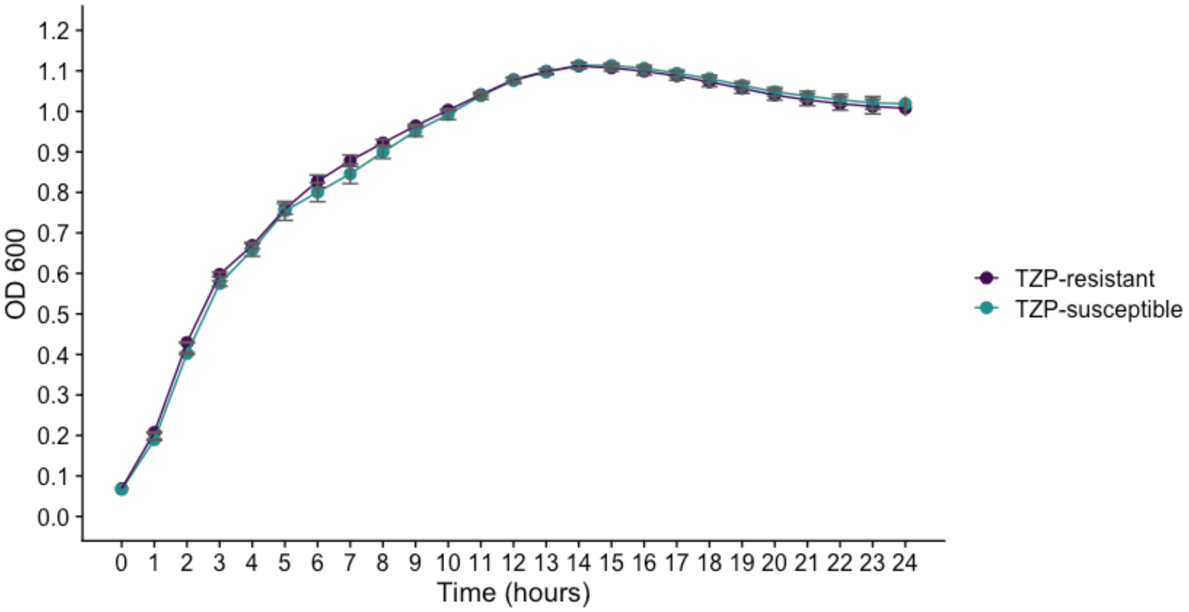


**Figure S3.** Growth curves of the TZP-S (green) and TZP-R (purple) *E. coli*. Overnight cultures grown from three TZP-S and three TZP-R colonies were equalised and then grown for 24 hours and 37 °C in LB broth. Optical density readings at 600 nm were obtained every hour. The points on the graph show mean values, error bars show the SEM.

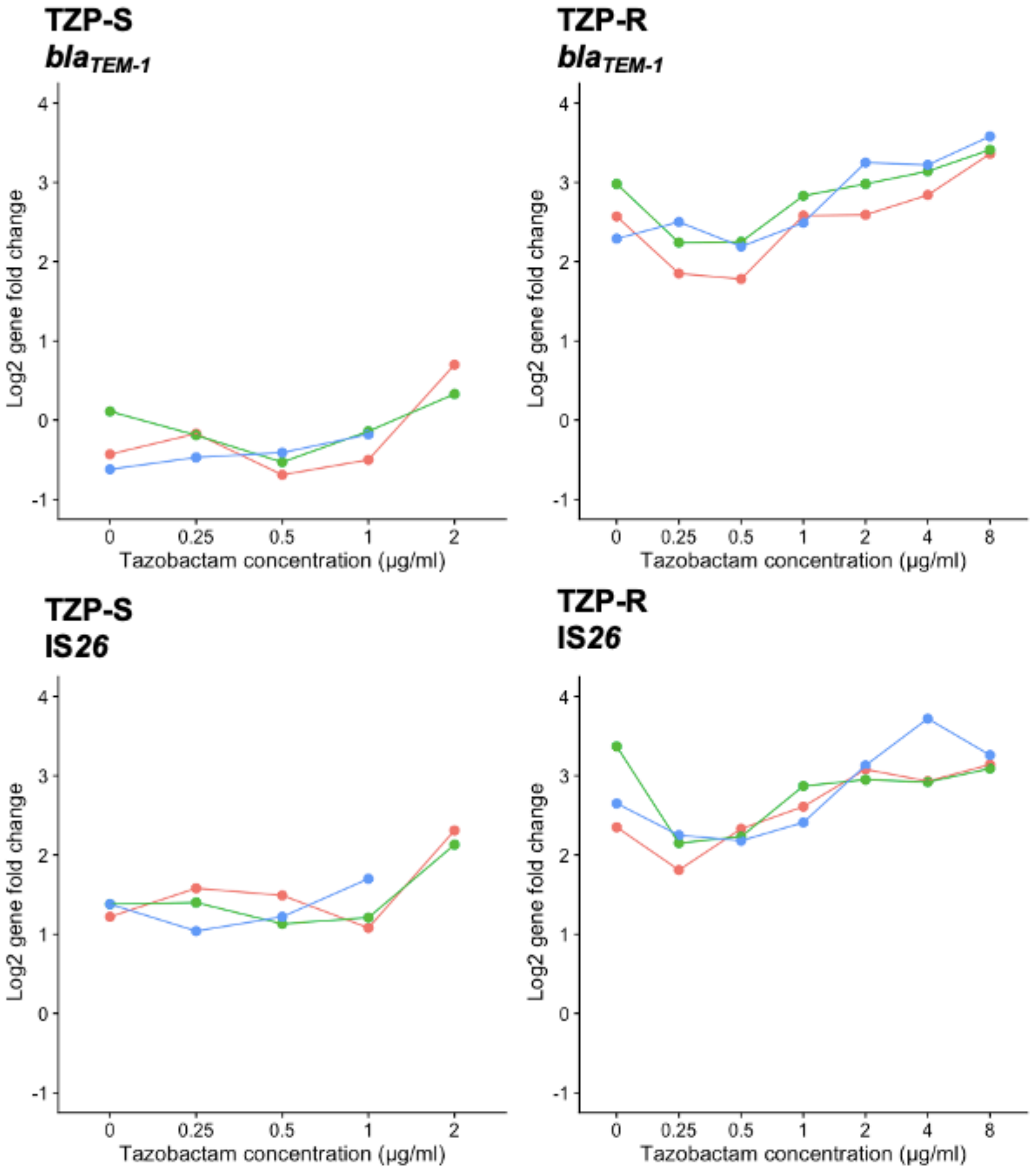


**Figure S4.** Gene copy number of *bla*$_{TEM-1}$ and *IS26* in the TZP-R and TZP-S isolate. following the culture of isolates in increasing concentration of tazobactam with piperacillin fixed at 8 μg/mL. Each colour on the plot represents a biological repeat. Copy number was calculated following normalisation against *uidA,* a single-copy housekeeping gene.

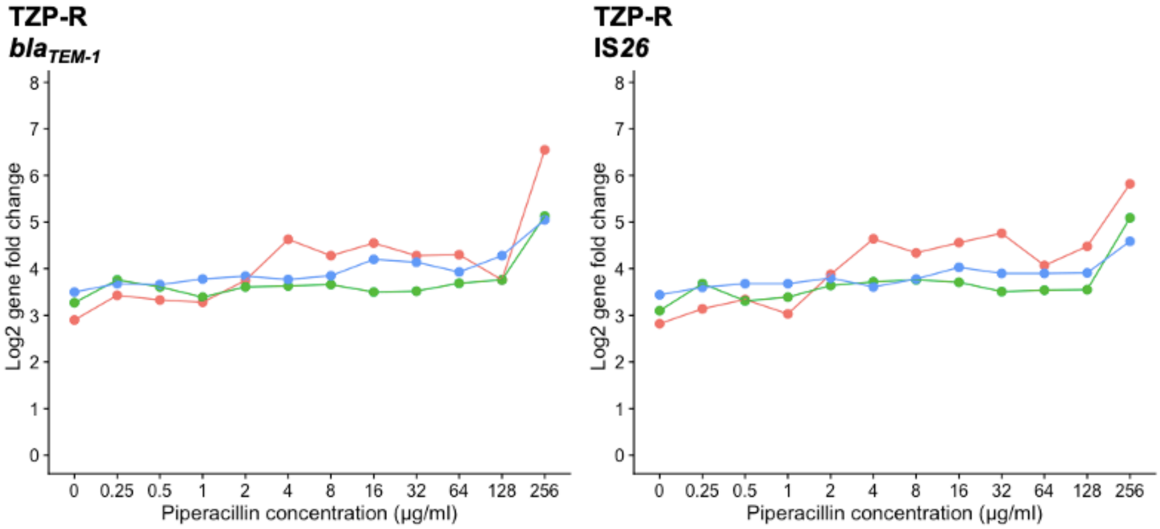


**Figure S5.** Gene copy number of *bla*$_{TEM-1}$ and *IS26* in the TZP-R isolate following culture in an increasing concentration of piperacillin with tazobactam fixed at 4 µg/mL. Each colour on the plot represents a biological repeat. Copy number was calculated following normalisation against *uidA,* a single-copy housekeeping gene.

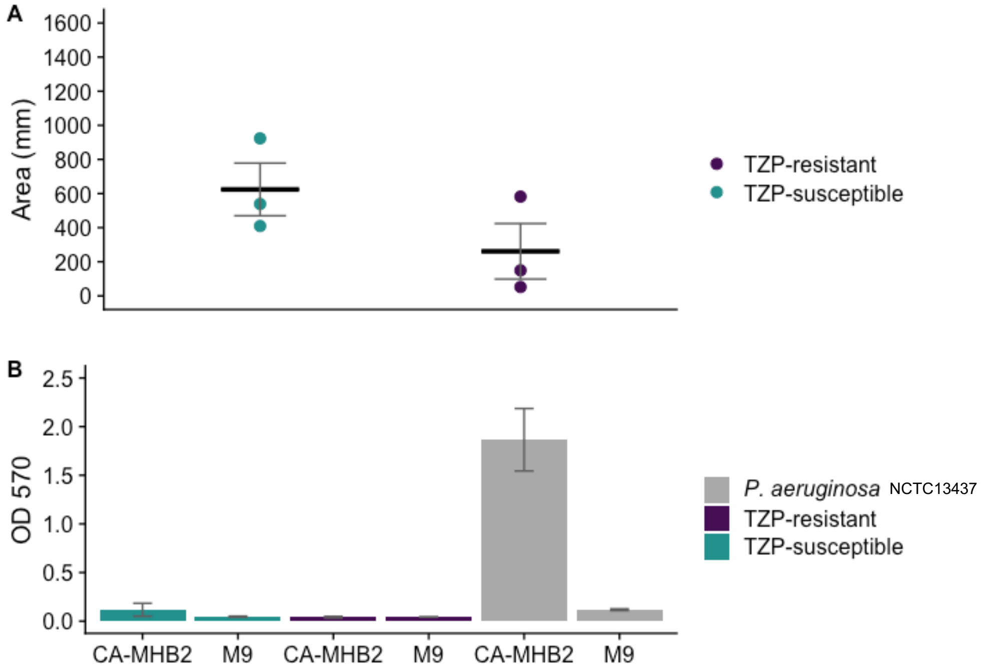


**Figure S6.** A motility assay **(A)** and biofilm assay **(B)** was used to detect the ability of the TZP-S (green) and TZP-R (purple) to initiate swimming motility and produce biofilm to enable adherence to polystyrene. Swimming motility was measured by calculating the area of the semi-solid (0.3%) agar that the bacteria spread through, from the inoculation point. A crystal violet assay was used for the quantitation of biofilm production (expressed as absorbance at $OD_{570}$ ). For each isolate, three biological repeats were performed, each containing three technical repeats. In plot **(A)** points show the mean for each group of three technical repeats, the horizontal black bar shows the mean of three biological repeats. In plot **(B)** the bars show the mean of the three biological repeats. In both plots error bars show the SEM.

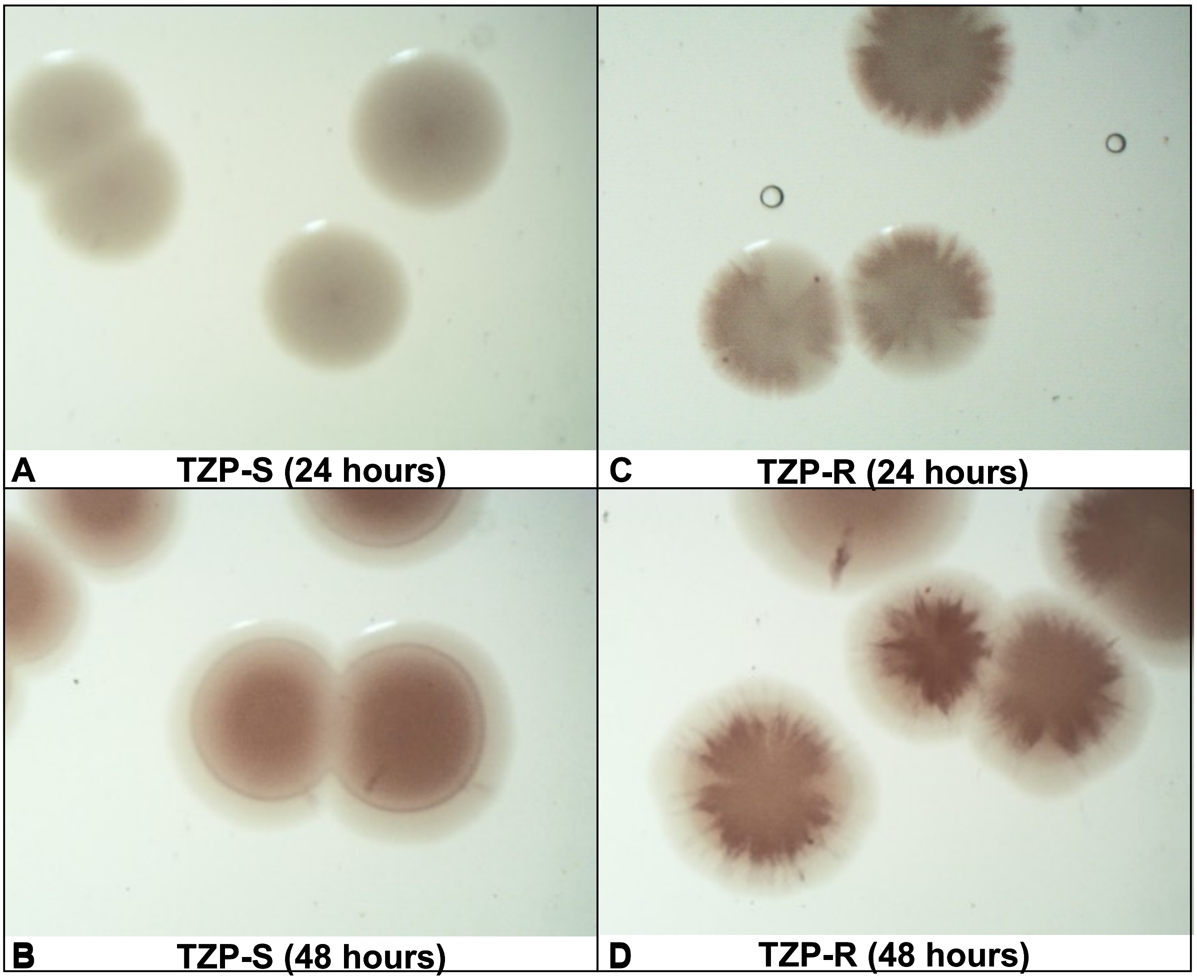


**Figure S7.** Colony morphotype and the production of extracellular matrix components were compared by growing the TZP-S **(A and B)** and TZP-R isolate **(C and D**) on LB agar supplemented with Brilliant Blue G-250 and Congo Red dye. Colonies were photographed after 24 hours and 48 hours of incubation at 37°C. For each isolate, three biological repeats, consisting of three technical repeats were performed.

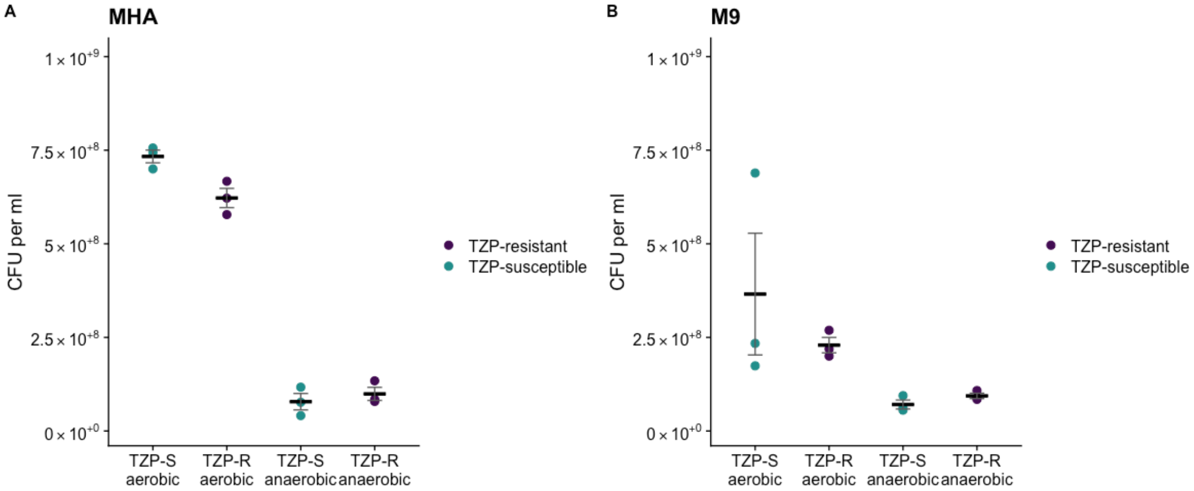


**Figure S8.** Growth assay using **(A)** MHA and **(B)** M9 agar, to compare CFU counts obtained when the TZP-S (green points) and TZP-R (purple points) isolate were grown in anaerobic conditions for 24 hours. For each isolate, three biological repeats, consisting of three technical repeats were performed. Each point is the mean of one group of three technical repeats. The black bar represents the mean on the biological repeats, error bars represent the SEM.

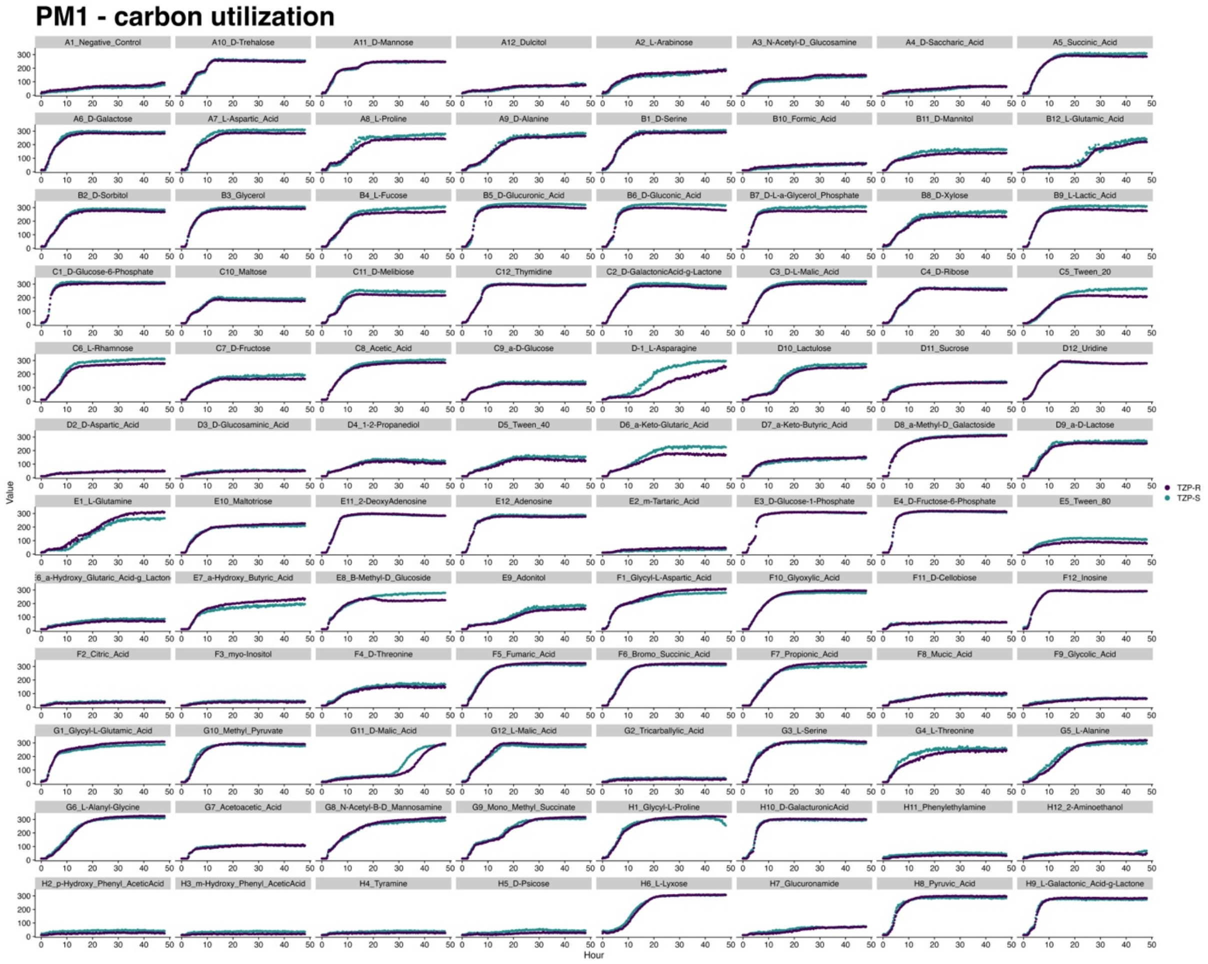


**Figure S9.** Average 48-hour growth curves for the TZP-S (green) TZP-R (purple) isolates, measured using the Biolog system and the phenotype microarray 1 (PM1) assay for carbon utilization. For each isolate, three biological replicates were performed and each point corresponds to the average of three replicates**.**

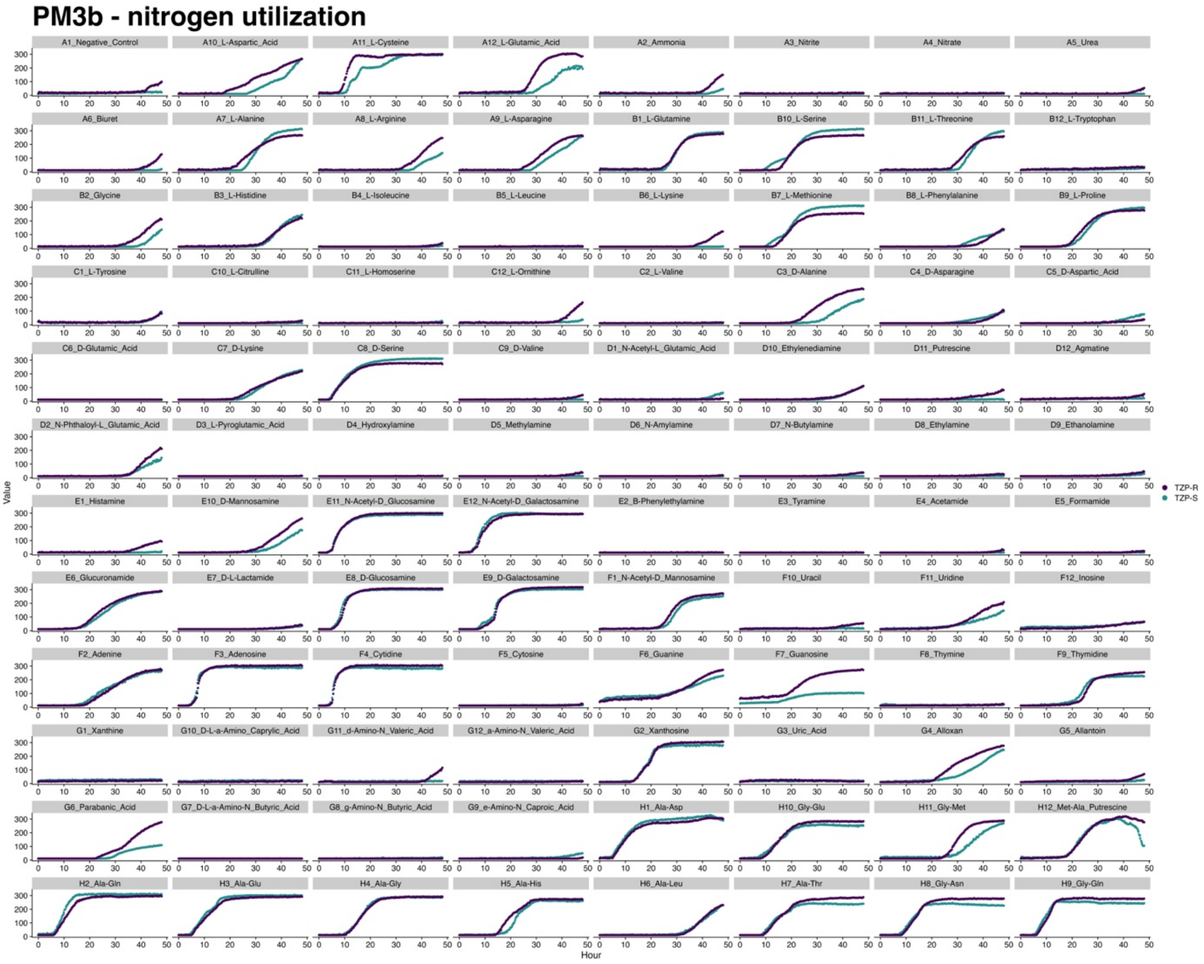


**Figure S10.** Average 48-hour growth curves for the TZP-S (green) TZP-R (purple) isolates, measured using the Biolog system and the phenotype microarray 3b (PM3b) assay for nitrogen utilization. For each isolate, three biological replicates were performed and each point corresponds to the average of three replicates**.**

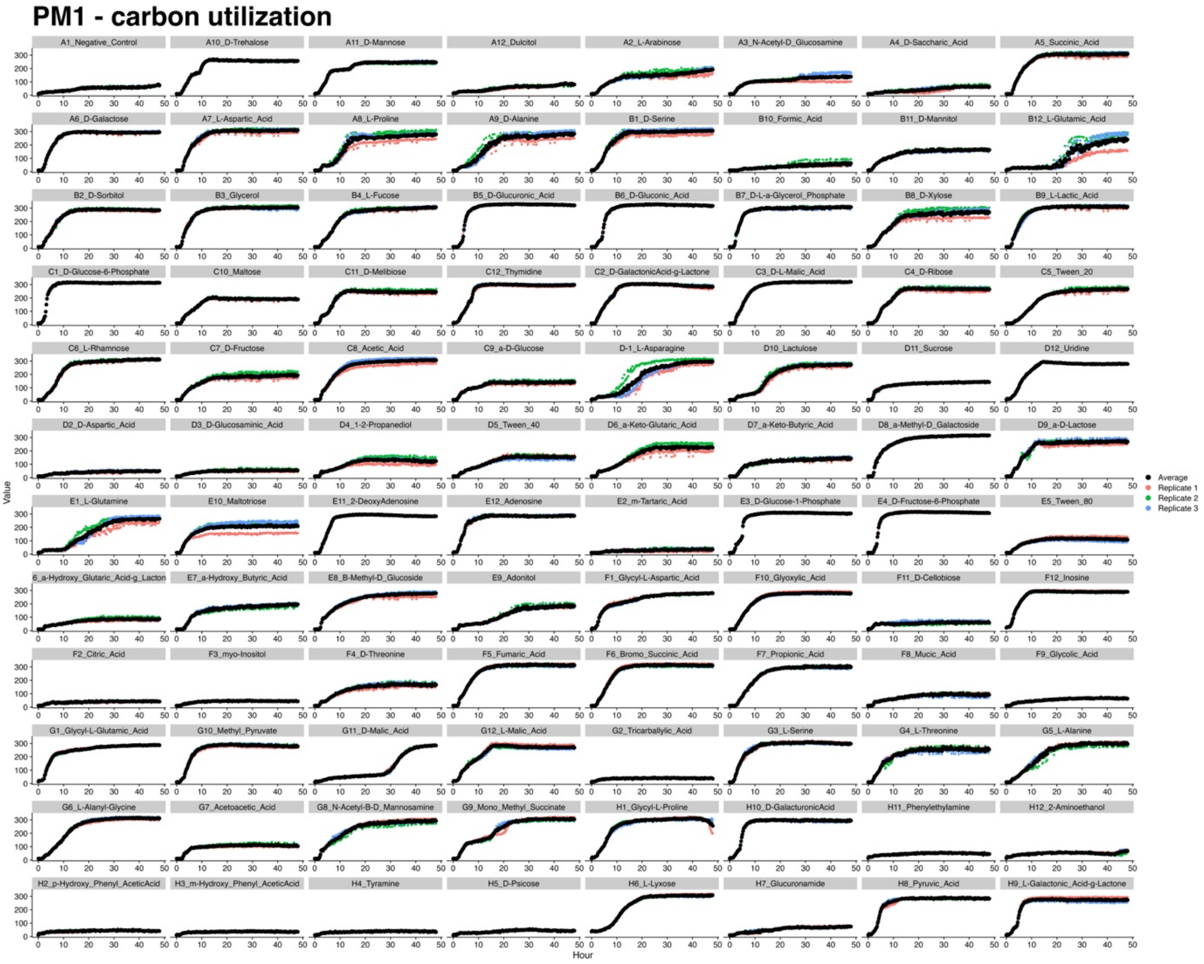


**Figure S11.** 48-hour growth curves for the TZP-S isolate, measured using the Biolog system and the phenotype microarray 1 (PM1) assay for carbon utilization. For each isolate, three biological replicates were performed. Each replicate is represented by a different colour (pink green or blue). Black points represent the average of the individual replicates.

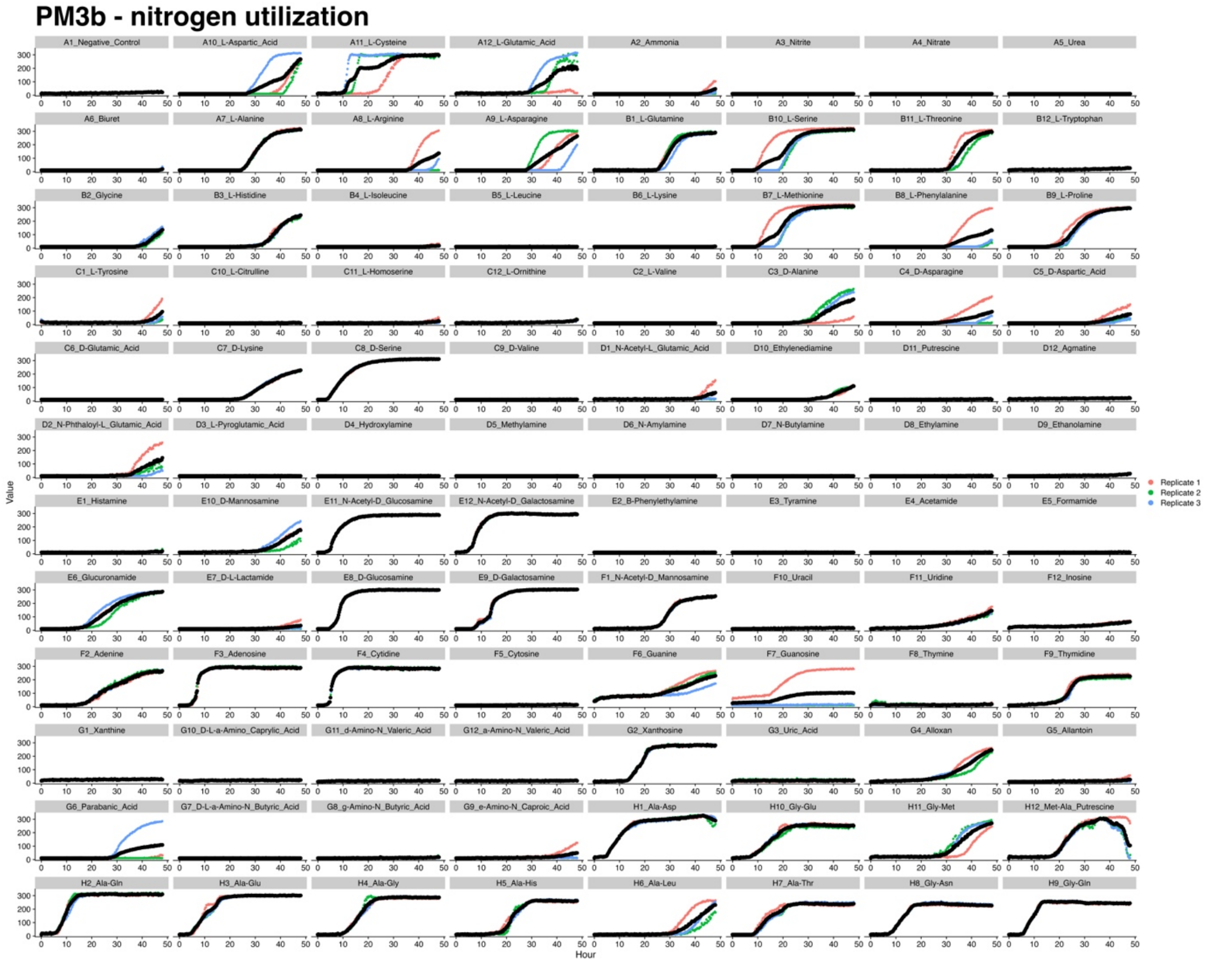


**Figure S12.** 48-hour growth curves for the TZP-S isolate, measured using the Biolog system and the phenotype microarray 3b (PM3b) assay for nitrogen utilization. For each isolate, three biological replicates were performed. Each replicate is represented by a different colour (pink green or blue). Black points represent the average of the individual replicates.

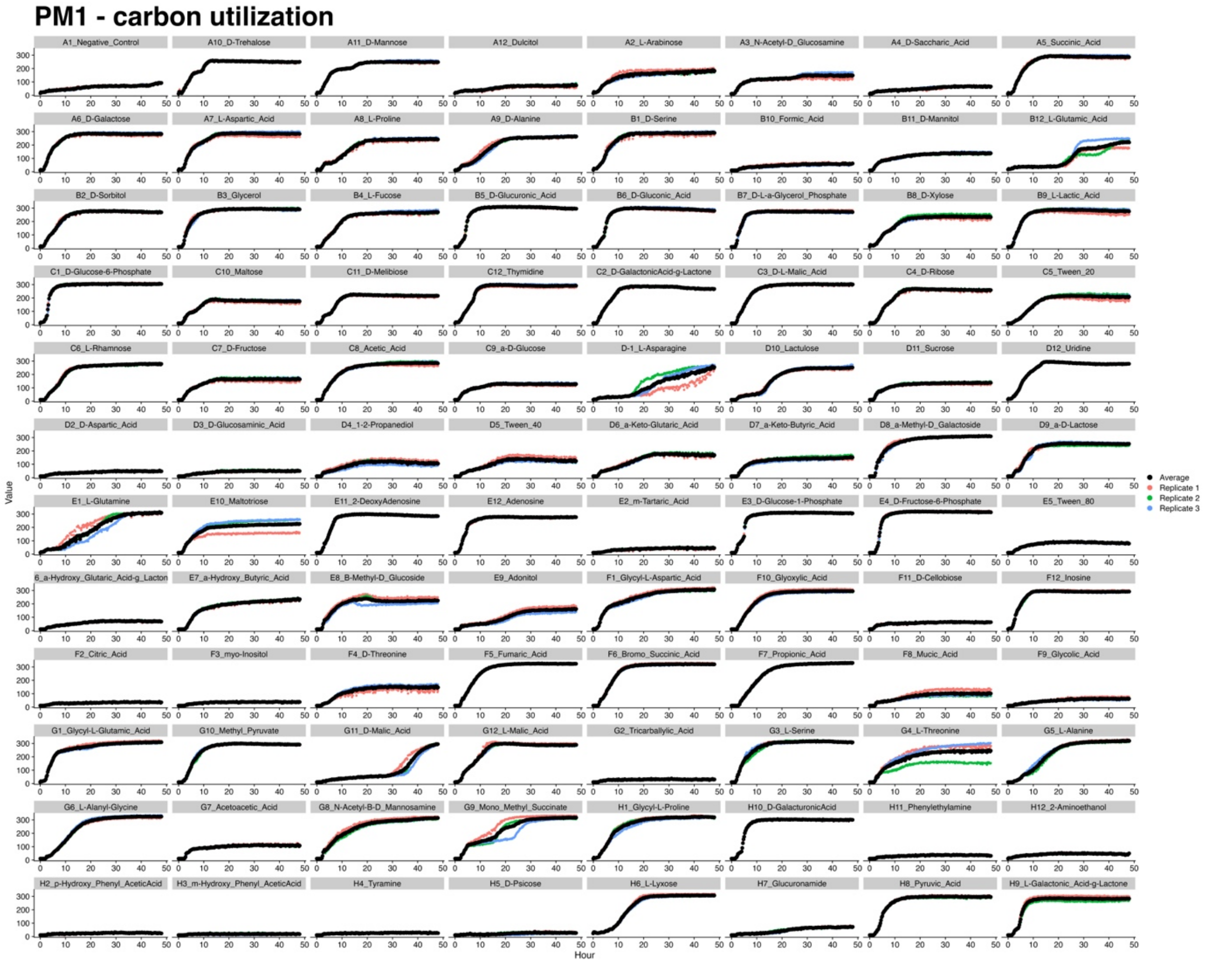


**Figure S13.** 48-hour growth curves for the TZP-R isolate, measured using the Biolog system and the phenotype microarray 1 (PM1) assay for carbon utilization. For each isolate, three biological replicates were performed. Each replicate is represented by a different colour (pink green or blue). Black points represent the average of the individual replicates.

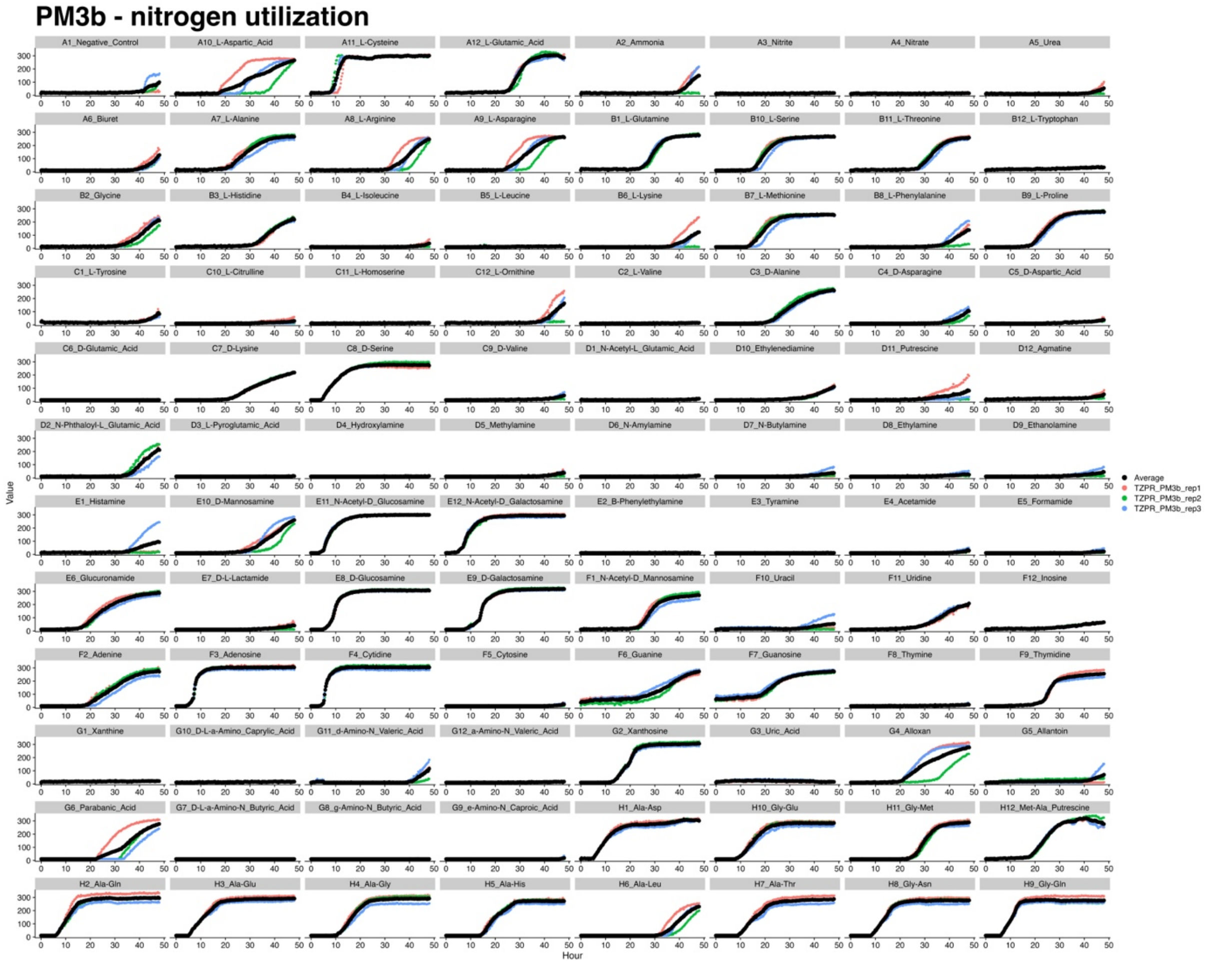


**Figure S14.** 48-hour growth curves for the TZP-R isolate, measured using the Biolog system and the phenotype microarray 3b (PM3b) assay for nitrogen utilization. For each isolate, three biological replicates were performed. Each replicate is represented by a different colour (pink green or blue). Black points represent the average of the individual replicates.
